# The effects of skin pigmentation on the accuracy of pulse oximetry in measuring oxygen saturation: a systematic review and meta-analysis

**DOI:** 10.1101/2022.02.16.22271062

**Authors:** Chunhu Shi, Mark Goodall, Jo Dumville, James Hill, Gill Norman, Oliver Hamer, Andrew Clegg, Caroline Leigh Watkins, George Georgiou, Alexander Hodkinson, Catherine Elizabeth Lightbody, Paul Dark, Nicky Cullum

## Abstract

**Background:** Pulse oximetry was widely used in hospitals and at home to monitor blood oxygen during the COVID-19 pandemic. There have been concerns regarding potential bias in pulse oximetry measurements for people with dark skin. We aimed to assess the effects of skin pigmentation on the accuracy of oxygen saturation measurement by pulse oximetry (SpO_2_) compared with the gold standard SaO_2_ measured by CO-oximetry.

**Methods:** We searched Ovid MEDLINE, Ovid Embase, and EBSCO CINAHL Plus (up to December 2021), as well as ClinicalTrials.gov and World Health Organization International Clinical Trials Registry Platform (up to August 2021). We identified studies comparing SpO_2_ values in any population, in any care setting, using any type of pulse oximeter, with SaO_2_ by standard CO-oximetry; and measuring the impact of skin pigmentation or ethnicity on pulse oximetry accuracy. We performed meta-analyses for mean bias (the primary outcome in this review) and its standard deviations (SDs) across studies included for each subgroup of level of skin pigmentation and ethnicity. We calculated accuracy root-mean-square (*A*_*rms*_) and 95% limits of agreement based on pooled mean bias and pooled SDs for each subgroup.

**Results:** We included 32 studies (6505 participants); 27/32 (84.38%) in hospitals and none in people’s homes. Findings of 14/32 studies (43.75%) were judged, via QUADAS-2, at high overall risk of bias. Fifteen studies measured skin pigmentation and 22 referred only to ethnicity. Compared with standard SaO_2_ measurement, pulse oximetry probably overestimates oxygen saturation in people with dark skin (pooled mean bias 1.11%; 95% confidence interval 0.29% to 1.93%) and people described as Black/African American (pooled mean bias 1.52%; 0.95% to 2.09%) (moderate- and low-certainty evidence). These results suggest that, for people with dark skin, pulse oximetry may overestimate blood oxygen saturation by around 1% on average compared with SaO_2_. The bias of pulse oximetry measurements for people with other levels of skin pigmentation, or those from the White/Caucasian group is more uncertain. The data do not suggest overestimation in people from other ethnic groups such as those described as Asian, Hispanic, or mixed ethnicity (pooled mean bias 0.31%, 0.09% to 0.54%), but this evidence is low certainty. Whilst the extent of mean bias is small or negligible for all the subgroups of population evaluated, the associated imprecision is unacceptably large (with the pooled SDs > 1%). Nevertheless, when the extents of measurement bias and precision are considered jointly in *A*_*rms*_, pulse oximetry measurements for all the subgroups appear acceptably accurate (with *A*_*rms*_ < 4%).

**Conclusions:** Low-certainty evidence suggests that pulse oximetry may overestimate oxygen saturation in people with dark skin and people whose ethnicity is reported as Black/African American, compared with SaO_2_, although the overestimation may be quite small in hospital settings. The clinical importance of any overestimation will depend on the particular clinical circumstance. Pulse oximetry measurements appear accurate but imprecise for all levels of skin pigmentation. The evidence relates to clinician-measured oximetry in health care environments and may not be reflected in home pulse oximetry where other factors may also influence accuracy.

**Author summary:** *Why was this study done?:*

- Pulse oximetry was widely used in hospital and at home to measure blood oxygen levels during the COVID-19 pandemic.
- There was uncertainty as to whether skin pigmentation affects the accuracy of pulse oximetry measurements.

*What did the researchers do and find?:*

- We assessed, via systematic review, the effects of skin pigmentation on the accuracy of pulse oximetry measurement (SpO_2_) compared with SaO_2_ measured by standard CO-oximetry.
- In people with dark skin, oxygen saturation measured in hospital by pulse oximetry may be overestimated by an average of 1% compared with gold standard SaO_2_, however, the evidence is of low certainty.
- The accuracy of pulse oximetry measured compared with standard SaO_2_ is quite uncertain for people with light or medium levels of skin pigmentation and for people from ethnic groups other than those described in papers as Black or African American.
- Pulse oximetry measurements appear to have acceptable overall accuracy (with *A*_*rms*_< 4%) for all subgroups of population evaluated whilst the variation of oximetry readings appear unacceptably wide (with the pooled SDs > 1%).

*What do these findings mean?:*

- Hospital measured pulse oximetry may overestimate oxygen saturation in people with dark skin compared with SaO_2_ by approximately 1%.
- The implications of this finding in different clinical scenarios will vary but could be clinically important. Impacts are likely to be at the thresholds of being diagnosed as having hypoxaemia where even a small SpO_2_ overestimation could lead to clinically important hypoxaemia remaining undetected and untreated.
- How these findings extrapolate to community and home care settings is unclear.

## Introduction

Blood oxygen saturation levels require monitoring for health reasons in a wide range of circumstances. Low blood oxygen saturation, if identified to be hypoxemia, requires medical intervention and has been linked to an increased risk of death.[1] The gold standard measure of blood oxygen saturation levels (SaO_2_) requires a sample of arterial blood and measurement using CO-oximetry. Pulse oximetry, measuring SpO_2_ as a proxy for SaO_2_ using a non-invasive and inexpensive device, is frequently used to detect low blood oxygen levels. Pulse oximetry has been widely used during the COVID-19 pandemic, including in non-clinical settings, to detect hypoxemia and inform decisions to escalate care.[2]

The current WHO COVID-19 management guideline recommends the “*use of pulse oximetry monitoring at home as part of a package of care*” for symptomatic people with COVID-19.[3] Many countries have specific guidance or services for home pulse oximetry in line with this recommendation.[2,4] In the UK, the NHS England COVID Oximetry@home service provided oximetry devices to people with COVID-19 for their home self-monitoring of oxygen levels. Guidance recommended that those with pulse oximetry readings of 92% or less “*attend A&E as quickly as possible or call 999 immediately*”.[2]

The reporting of possible bias in pulse oximetry measurement, including due to skin pigmentation, raised a growing concern about the accuracy of oxygen self-monitoring.[5] Measurement bias could have serious clinical implications including the delay of urgent medical care.[6]. A recent US study analysed retrospective cohort data from more than 10,000 people, comparing where a diagnosis of *occult hypoxemia* (an SaO_2_ of less than 88%) was missed by pulse oximetry.[7] Results showed people described as Black had ‘*nearly three times the frequency of occult hypoxemia that was not detected by pulse oximetry*’ as those described as White.[7] In November 2021, the UK Health Secretary ordered a review into racial bias in medical equipment, including pulse oximeters.

It is an important time to consider the current evidence base for the impact of skin pigmentation on the accuracy of pulse oximetry compared with the gold standard measure of SaO_2_. The only current relevant systematic review, published in 1995, included three studies that explicitly considered the impact of skin pigmentation on pulse oximetry accuracy.[8] The review concluded that pulse oximeters may overestimate blood oxygen saturation in people with high levels of skin pigmentation.[8] The recent rapid review by the NHS Race and Health Observatory came to similar conclusions but used a non-systematic review process, e.g., having no comprehensive searches, critical appraisal of included studies and meta-analysis.[6] Our objective was therefore to conduct a systematic review of relevant evidence for the influence of skin pigmentation on the accuracy of oxygen saturation measurement by pulse oximetry (SpO_2_) compared with SaO_2_ measured by standard CO-oximetry.

## Methods

We used Cochrane methodologies to conduct this review,[9] and report it in accordance with the Preferred Reporting Items for Systematic Reviews and Meta-Analyses (PRISMA) statement.[10]

### Protocol and registration

We registered the protocol with the Open Science Framework (available here https://osf.io/gm7ty).[11]

### Eligibility criteria

We included any prospective or retrospective methods-comparison study, completed or ongoing, including full publications and conference abstracts.

We included studies that compared SpO_2_ values in any population, in any care setting, measured using any type of commercially available pulse oximeter, with SaO_2_ measured by standard CO-oximetry (in the same individuals).[12] We excluded studies that used: (1) prototype pulse oximetry devices; (2) pulse oximeters that require high-skilled specialists to operate (such as intra-partum pulse oximetry devices); and (3) pulse oximeters used for measuring venous blood oxygen saturation, as our focus was evaluating commercial pulse oximetry devices that could be potentially used at home for arterial oxygen saturation measurement.

We included studies investigating the accuracy of pulse oximetry based on both the level of skin pigmentation and ethnic group. We considered ethnicity as a sub-optimal but relevant proxy for level of skin pigmentation in relation to the accuracy of the biological reading of oxygen saturation.

For level of skin pigmentation measurement, we included studies that used a standardised measure of level of skin pigmentation such as the Fitzpatrick scale,[13] and studies that used an unstandardised or qualitative judgement of skin pigmentation levels such as so called ‘light’ or ‘dark’ level of skin pigmentation. The originally reported terms of skin pigmentation were mapped into ‘low’, ‘medium’ or ‘high’ pigmentation categories, without any further re-classification. For example, if a study classified skin as ‘light’ and ‘dark’, then the light skin group was mapped into the low pigmentation category, and the dark skin group into the high pigmentation category. For ethnicity, we included studies that grouped participants based on any ethnicity classification. We interpreted terms such as Black and White as describing ethnicity and only indirect indicators of skin pigmentation.[14]

Where study reports were inadequate to support inclusion decisions, we contacted authors for more information. We excluded studies where no response was received from contacting authors.

We focus here on reporting the comparative accuracy of pulse oximetry-produced SpO_2_ in relation to SaO_2_ measured by gold standard CO-oximetry. We therefore excluded studies that reported diagnostic test accuracy measures and those with inappropriate comparators, including: use of a reference pulse oximetry as the comparator, use of incorrect reference values of oxygen saturation e.g. arterial oxygen pressure (PaO_2_), calculated SaO_2_, fractional saturation (%O_2_Hb or FO_2_Hb).[12,15]

Following the British Standards Institution 2019 standards for pulse oximetry,[12] we included data on the overall accuracy (agreement between a test result and an accepted reference value reported as accuracy root-mean-square, *A*_*rms*_), mean bias and precision (and/or the limits of agreement for the SpO_2_ and SaO_2_ comparison) (Supporting Information Table S1). The overall accuracy (*A*_*rms*_) is a combination of mean bias and precision in a single measure.[12]. Mean bias is calculated as the mean difference between two measures (in this case SpO_2_ – SaO_2_).[12] A larger mean difference indicates a larger bias value and values greater than 0 indicate overestimation with pulse oximetry.[12] Precision is commonly reported as the standard deviation of between-test mean difference with a larger standard deviation indicating less precision.[12]

The British Standards Institution standards for pulse oximetry gives *A*_*rms*_ primacy, as it details general accuracy and thus the suitability of the machine for its purpose. The value is a root mean square deviation calculation, and the relevance of this measure to clinical decision-making is not intuitive. Because of this we present mean bias as the review’s primary outcome. This mean difference between ‘true’ blood oxygen saturation levels and those measured by pulse oximetry can more clearly indicate how clinical decisions referring to threshold values (e.g., admission to hospital with a pulse oximetry reading of 92% or lower) could be impacted by bias.

### Information sources and literature searches

In August 2021, we searched Ovid MEDLINE and Epub Ahead of Print, In-Process, In-Data-Review & Other Non-Indexed Citations and Daily (1946), Ovid Embase (1974), and EBSCO CINAHL Plus (Cumulative Index to Nursing and Allied Health Literature; 1937) to identify only English language reports of relevant studies given the limited time and resources available. Searches were updated in December 2021. There were no restrictions with respect to date of publication or study setting. See Supporting Information Box S1 for the search strategy used for Ovid MEDLINE.

In August 2021, we searched the ClinicalTrials.gov and World Health Organization International Clinical Trials Registry Platform for ongoing studies; and the reference lists of retrieved included studies, relevant systematic reviews, and guideline reports. We also contacted authors of key abstracts to request further information about their studies.

### Study selection

Two reviewers (CS and MG, or JH, OH) independently assessed titles and abstracts of the search results for relevance, with records stored and screened in the Abstrackr software.[16] Two reviewers (CS and MG or JH) independently inspected the full texts of all potentially eligible studies. The two reviewers resolved disagreements through discussion, involving a third reviewer where necessary.

### Data extraction

One reviewer (CS, or OH or JH) independently extracted data from included studies, which were checked by another reviewer (JH, MG, OH, GN). We resolved any disagreements through discussion. See Supporting Information Box 2 for data items we extracted.

We contacted study authors to clarify methods and data, where necessary. We transformed data into a format needed for analyses when required, e.g. from reported 95% limits of agreement or confidence interval (CI) to standard deviation (SD) using Wan’s method.[17]

### Risk of bias assessment

There is no standard risk of bias tool for methods-comparison studies. We therefore chose QUADAS-2 for risk of bias assessment;[18] tailoring it as required, by adding and omitting signalling questions, see Supporting Information Box 3 for further information.

One reviewer (CS, or OH or JH) independently assessed the risk of bias for the included studies, checked by another reviewer (GN, MG, JH, OH) with any discrepancies resolved via discussion.

## Data synthesis and analysis

We summarised the included studies narratively and quantitatively; pooling evidence on *A*_*rms*,_ mean bias, precision (SD), and 95% limits of agreement. As specified in our protocol, we analysed data separately for studies which reported level of skin pigmentation and ethnicity. When pooling data for mean bias and its SD across studies, we used the correlated hierarchical effects model with small-sample corrections under the Robust Variance Estimation (RVE) framework (Supporting Information Box 4). The approach enabled us to include single-measure design study data, together with multiple dependent effect size estimates (i.e. mean bias and SD) of a repeated-measures design study in meta-analysis even when the dependence structure is unknown.[19–21] We used Tau^2^ (95% CI), I^2^ and the Q statistic and the related Chi^2^ test to fully assess heterogeneity in meta-analysis.

There is no established approach to pooling data for *A*_*rms*_ and 95% limits of agreement across studies directly. In this review, we used the pooled mean bias and the pooled SDs produced by related meta-analyses, and followed the British Standard Institute methods to calculate the *A*_*rms*_ [12] and Tipton and Shuster’s methods to calculate the population 95% limits of agreement.[20,22]

Within the R open-source software environment (version 4.1.2; R Foundation for Statistical Computing),[23] we performed RVE meta-analyses using functions available in packages of *clubSandwich*,[24] and *metafor*,[25] and used the *forestplot* package to produce forest plots that graphically present pooled – or originally reported when data pooling was impossible – results of *A*_*rms*_, mean bias (SD), and 95% limits of agreement.[26] Supporting Information Box 4 presents generic R codes used for meta-analysed. All codes and analyses were checked by our statistician co-author (AH).

When meta-analysis was not appropriate, we synthesised relevant evidence following the Synthesis Without Meta-analysis in systematic reviews (SWiM) reporting guideline.[27]

One reviewer (CS) assessed the certainty of evidence on mean bias by referring to the GRADE approach developed for the test accuracy topic, checked by another reviewer (GN).[28,29] Using this approach the certainty of mean bias findings could be assessed as at high, moderate, low or very low certainty. In interpreting review findings, we used the British Standards Institution-recommended thresholds to judge the accuracy of pulse oximetry, viz. for pulse oximetry to be accurate by British Standards Institution standards, the overall accuracy *A*_*rms*_ should be within 4% over the range of 70% to 100% SaO_2_. For the mean bias (and precision), SpO_2_ measures should be within +/-2% of CO-oximetry measures and the variation for repeated SpO_2_ measures should be within one SD of the mean bias.[12] With the mean bias as the primary outcome, any pooled mean bias of > 0% would suggest a risk of hypoxemia. Given pulse oximeter devices commonly present integers in percentage, we rounded pooled estimates to be integers when interpreting the related findings such as rounding mean bias values within +/-0.50% to 0%.

We analysed data on pulse oximeters of different brands/manufacturers separately where possible, or presented originally reported data where meta-analysis was not appropriate.

We undertook pre-planned sensitivity analyses to assess the robustness of meta-analysis, (1) excluding studies in which all participants had the same level of skin pigmentation or were from the same ethnicity, (2) excluding studies that have no data directly available for a meta-analysis unless data are transformed, and (3) excluding studies at high overall risk of bias. We also undertook post-hoc sensitivity analysis to assess the impact of excluding studies that used descriptors of ethnicity to indicate levels of skin pigmentation. This is because those study data might be systematically different from the others that used standardised skin pigmentation measurement methods such as the Fitzpatrick scale.

We assessed publication bias following a qualitative approach without using funnel plots or Egger’s tests, given existing methods were not considered appropriate for the case of this review.[30]

## Results

### Search results

Following a search in August 2021 and an update search in December 2021, we assessed titles and abstracts of 9920 records identified from electronic databases and 152 from trial registries. We also assessed 14 records identified by screening the reference lists of included studies and existing reviews. After assessing full texts of 382 potentially relevant records, we identified 33 publications of 32 studies – published between 1985 and 2021 – as eligible for inclusion in this review (Figure 1).[31–63] We also identified one ongoing study from electronic searches.[64] By contacting authors we received raw or study-level summary data for two studies.[39,51]

**Figure 1.**
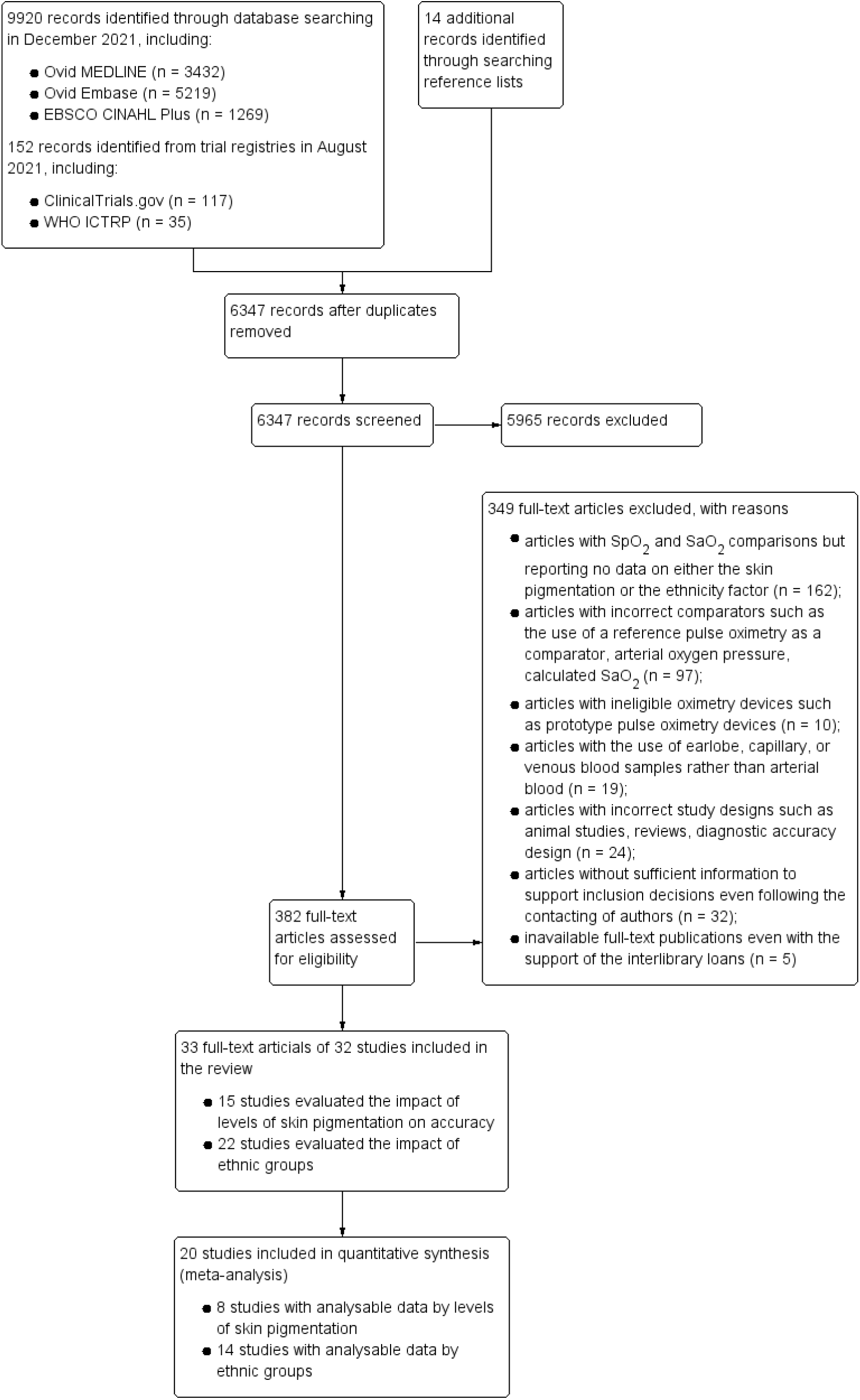
The study selection flowchart.

### Characteristics of included studies

Table 1 summarises included studies (Supporting Information Table S2 presents a more detailed overview).

**Table 1.** Summary characteristics of the included studies

| Items | Summary statistics (n (%)) |
| --- | --- |
| Study designs (32 studies) |  |
| *Prospective design* | 29 (90.62%) |
| *Retrospective design* | 3 (9.38%) |
| Repeated-measures design (32 studies) |  |
| *Yes (that is, more than one SpO<sub>2</sub>-SaO<sub>2</sub> data pair collected per person)* | 21 (65.62%) |
| *No* | 11 (34.38%) |
| Care settings (32 studies) |  |
| *Hospital care settings* | 27 (84.38%) |
| *Laboratory setting\** | 5 (15.62%) |
| Types of participants (32 studies) |  |
| *Children* | 7 studies (21.88%), with 1608 participants involving:<br>• a current critical illness,[33,36,61]<br>• hypoxemic conditions and/or cyanotic congenital heart disease,[40,42,43,55] |
| *Adults* | 25 (78.12%), with 4897 participants involving a variety of health conditions:<br>• healthy volunteers,[34,39,57,59,63]<br>• critical illnesses or conditions needing intensive care unit admission and/or mechanical ventilation,[35,37,44-48,56]<br>• pulmonary/respiratory conditions including COVID-19,[38,49,53,54,60,62]<br>• cirrhosis,[31]<br>• chronic rheumatic heart disease,[58]<br>• postoperative hypothermia,[41]<br>• hospitalised patients in general,[32,51]<br>• adults under the need of a long-term home oxygen therapy[50] |
| Sample sizes (32 studies) | Median 50 (range: 6 to 1562) |
| Age (32 studies) |  |
| Mean or median specified (n = 23) | Median 56.40 years (range: 4 days to 69 years) |
| Factors related to skin pigmentation (32 studies) |  |
| *Levels of skin pigmentation* | 15 studies (46.88%), with 1800 participants |
| *Descriptors of ethnicity* | 22 studies (68.88%), with 4910 participants |

Note: \* The studies at the laboratory setting (n = 5) only recruited healthy volunteers.

The 32 studies (6505 participants) reported SpO_2_-SaO_2_ comparison evaluations of 54 different pulse oximeters from 26 manufacturers. As the gold standard comparator measure, SaO_2_ was directly measured using either one of six generic (or 14 specific) types of CO-oximetry methods, or one of two combinations of CO-oximetry methods (Supporting Information Table S3).

### Assessment of risk of bias and applicability

Using QUADAS-2, we considered 14/32 studies (43.75%) to be at unclear risk of bias for all four domains or high risk of bias for at least one domain,[34,38,44,47,48,53,54,56–61,63] and the remaining 18 (56.25%) to be at low risk of bias for at least one of the four domains (Figure 2).

**Figure 2.**
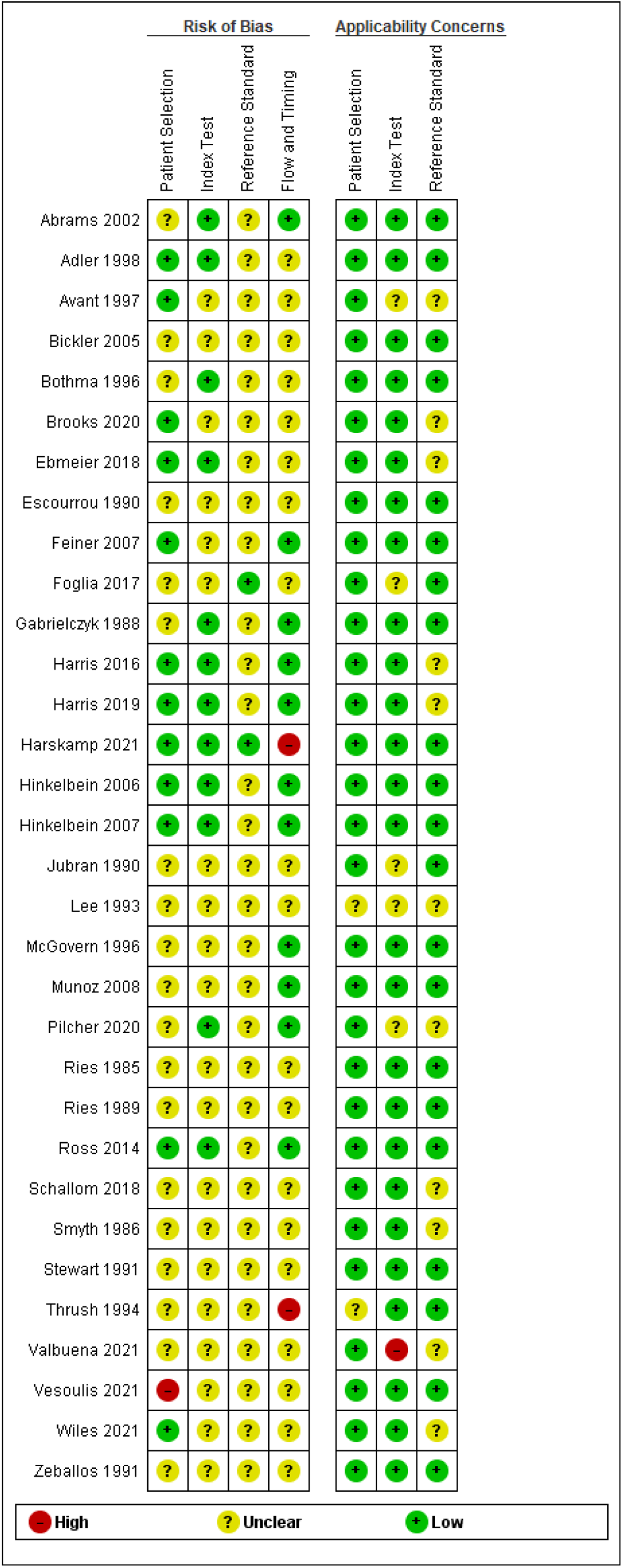
Risk of bias assessment results.

Key limitations in downgrading risk of bias were: (1) for the patient selection domain, where specific sub-populations were inappropriately excluded from a study[61] or there was no explicit exclusion or even inclusion criteria (19 studies); (2) for index test and reference standard domains, where there was no blinding information for either pulse oximetry SpO_2_ measurements (20 studies) or CO-oximeter SaO_2_ readings (30 studies); and (3) for the flow and timing domain, where there were sub-optimally long time intervals between SpO_2_ readings and the arterial blood sampling for SaO_2_ measurement,[44] or the exclusion of participants (3/25, 12.00%) from study data analysis without rationale.[59]

We judged the applicability concern as high for one study,[60] moderate for 13 studies,[33,36,37,40,42,43,47,48,51,56,57,59,62] and low in terms of all three applicability considerations for the remaining 18 studies. Applicability concerns largely resulted from the lack of specifications on pulse oximeters evaluated, CO-oximeter devices used, and/or arterial blood sampling procedures, thus SpO_2_ or SaO_2_ could not be reproduced.

### Association between level of skin pigmentation and pulse oximetry accuracy

Fifteen of the 32 studies (1800 participants) reported by level of skin pigmentation.[32,34,35,37–44,51–54,63] Eight of these studies (1297 participants) had available data and were included in the meta-analyses:[32,34,35,37,39,40,51,52,63] Supporting Information Table S4 presents the mapping of originally reported terms of skin pigmentation into ‘low’, ‘medium’ or ‘high’ pigmentation categories. Table 2 presents pooled association data (further data detail along with GRADE assessment details are found in Supporting Information Figures S1-S3 and Table S5).

**Table 2.** Result summaries of meta-analysis for levels of skin pigmentation and ethnic groups

| Subgroup categories | No. of studies (evaluations) | Sample size (data pairs) | Calculated $A_{rms}$, % | Pooled mean bias (95% CI), % | Pooled SD (95% CI), % | Calculated 95% limits of agreement, % | Overall $I^2$ (between-studies and within-study heterogeneity) in mean bias data pooling |
| --- | --- | --- | --- | --- | --- | --- | --- |
| **High (dark) skin pigmentation** |  |  |  |  |  |  |  |
| Primary analysis | 8 (24) | 221 (3270) | 1.88 | 1.11 (0.29 to 1.93) | 1.52 (1.30 to 1.79) | -1.87 to 4.09 | 98.03% (0% and 98.03%) |
| Sensitivity analysis, excluding studies with high overall risk of bias | 6 (15) | 177 (2691) | 1.75 | 0.87 (-0.46 to 2.19) | 1.52 (1.20 to 1.93) | -2.11 to 3.84 | — |
| Sensitivity analysis, excluding studies with no use of standardised scales for measuring skin pigmentation | 5 (9) | 160 (474) | 1.79 | 0.89 (-1.37 to 3.14) | 1.55 (1.12 to 2.15) | -2.16 to 3.93 | — |
| Sensitivity analysis, excluding studies with participants of high skin pigmentation alone | 6 (14) | 88 (2738) | 1.95 | 1.12 (-0.16 to 2.39) | 1.60 (1.26 to 2.03) | -2.02 to 4.26 | — |
| **Medium pigmentation** |  |  |  |  |  |  |  |
| Primary analysis | 4 (10) | 406 (1323) | 1.58 | -0.58 (-2.25 to 1.09) | 1.47 (1.08 to 2.00) | -3.46 to 2.30 | 92.65% (82.39% and 10.25%) |
| Sensitivity analysis, excluding studies with no use of standardised scales for measuring skin pigmentation | 3 (4) | 399 (399) | 1.66 | -0.79 (-3.03 to 1.45) | 1.46 (1.00 to 2.14) | -3.66 to 2.08 | — |
| **Low (light) skin pigmentation** |  |  |  |  |  |  |  |
| Primary analysis | 6 (15) | 670 (2865) | 1.53 | -0.35 (-1.36 to 0.67) | 1.49 (1.23 to 1.81) | -3.27 to 2.58 | 92.73% (22.42% and 70.31%) |
| Sensitivity analysis, excluding studies with high overall risk of bias | 5 (12) | 660 (2245) | 1.57 | -0.47 (-1.77 to 0.83) | 1.50 (1.21 to 1.87) | -3.42 to 2.47 | – |
| Sensitivity analysis (scale) | 4 (6) | 648 (667) | 1.62 | -0.54 (-2.52 to 1.43) | 1.53 (1.18 to 1.98) | -3.53 to 2.45 | – |
| **Participants described as Black/African American** |  |  |  |  |  |  |  |
| Primary analysis | 9 (22) | 459 (5753) | 2.27 | 1.52 (0.95 to 2.09) | 1.68 (1.32 to 2.14) | -1.78 to 4.82 | 96.39% (0% and 96.39%) |
| Sensitivity analysis, excluding studies with high overall risk of bias | 4 (10) | 67 (2892) | 1.99 | 1.47 (-0.21 to 3.16) | 1.35 (1.08 to 1.69) | -1.17 to 4.11 | – |
| Sensitivity analysis, excluding studies with data transformation | 7 (20) | 316 (3110) | 2.26 | 1.55 (0.85 to 2.25) | 1.64 (1.28 to 2.11) | -1.67 to 4.77 | – |
| Sensitivity analysis, excluding studies with Black/African American populations alone | 8 (16) | 426 (5621) | 2.35 | 1.63 (0.77 to 2.49) | 1.69 (1.25 to 2.28) | -1.68 to 4.94 | – |
| **Participants from other ethnic groups, described as Asian, Hispanic or mixed ethnicity** |  |  |  |  |  |  |  |
| Primary analysis | 3 (9) | 522 (2646) | 1.58 | 0.31 (0.09 to 0.54) | 1.55 (0.53 to 4.53) | -2.72 to 3.35 | 47.95% (0% and 47.95%) |
| Sensitivity analysis, excluding studies with high overall risk of bias | 2 (7) | 41 (2165) | 1.23 | 0.36 (-0.24 to 0.95) | 1.18 (0.14 to 9.78) | -1.95 to 2.66 | – |
| Sensitivity analysis, excluding studies with data transformation | 2 (8) | 488 (1405) | 2.07 | 0.30 (-0.80 to 1.40) | 2.04 (0.32 to 12.81) | -3.70 to 4.31 | – |
| **White/Caucasian** |  |  |  |  |  |  |  |
| Primary analysis | 13 (48) | 2195 (12870) | 1.64 | 0.55 (-0.21 to 1.31) | 1.55 (1.31 to 1.82) | -2.48 to 3.58 | 94.39% (69.92% and 24.47%) |
| Sensitivity analysis, excluding studies with high overall risk of bias | 8 (26) | 1293 (7770) | 1.42 | 0.36 (-0.88 to 1.61) | 1.38 (1.19 to 1.59) | -2.33 to 3.06 | – |
| Sensitivity analysis, excluding studies with data transformation | 10 (45) | 1044 (5485) | 1.67 | 0.63 (-0.38 to 1.63) | 1.54 (1.32 to 1.80) | -2.40 to 3.65 | – |
| Sensitivity analysis, excluding studies with White/Caucasian populations alone | 8 (16) | 1246 (9791) | 1.84 | 0.98 (-0.08 to 2.05) | 1.56 (1.16 to 2.10) | -2.07 to 4.04 | – |

Notes. Calculated accuracy root-mean-square ( $A_{rms}$ ) was derived based on the pooled mean bias and the pooled SD from related meta-analyses (Supporting Information Box S4). According to the British Standards Institution standards,[12] pulse oximetry with an $A_{rms}$ larger than 4% is considered inaccurate.
The pooled mean bias was produced by meta-analysis of data on mean SpO<sub>2</sub>–SaO<sub>2</sub> bias. Similarly, the pooled SD was from meta-analysis of all SD data reported in included studies. According to a commonly used criteria (mean bias of +/- 2% (one SD)), [12] an accurate pulse oximetry should produce SpO<sub>2</sub> measures that are within +/- 2% of CO-oximetry measures and the variation of bias values for repeated SpO<sub>2</sub> measures should be within one SD.
The 95% limits of agreement was calculated using the method described by Bland and Altman,[22] based on the pooled mean bias, and the pooled SD (Supporting Information Box S4).
Full results of heterogeneity tests are in Supporting Information Figures S1-S3 and S5-S7.

People with high levels of pigmentation probably have overestimated oxygen saturation readings from hospital-based pulse oximetry compared with standard SaO_2_ readings (8 studies, 24 comparisons, 3270 SpO_2_-SaO_2_ pairs from 221 participants): pooled mean bias 1.11% (95% CI 0.29% to 1.93%), moderate-certainty evidence. This means that, on average, pulse oximetry probably overestimates blood oxygen saturation by approximately 1% in this group, but the overestimation may be as low as 0.29% or almost 2%. The evidence for people with medium skin pigmentation is uncertain, meaning further research is likely to alter findings. The evidence for people with light skin pigmentation does not suggest clinically important systematic bias (pooled mean bias −0.35, 95% CI −1.36 to 0.67), but it is of low certainty.

For all the levels of skin pigmentation, the *A*_*rms*_ values are around 2% or lower (95% CI non-estimable), and the pooled SD values are around 1.50% on average (Table 2). These data mean that, for people with any level of skin pigmentation, about 68% of their pulse oximetry readings would be within ±2% of the CO-oximetry readings, with one SD indicating a variation around the mean bias of minus 1.50% to plus 1.50%.

We tested the sensitivity of the findings: A_*rms*_ and SD values were generally consistent but there was increased uncertainty for mean bias findings. Supporting Information Figure S4 presents evidence for different types of pulse oximeters evaluated: overall, most devices slightly overestimated oxygen saturation in people with high levels of skin pigmentation, with imprecision around estimates.

Seven of the 15 studies (with 503 participants) were not included in meta-analysis (Supporting Information Table S6):[38,41–44,53,54] six compared differences in mean bias between pigmentation levels, rather than reporting mean bias by levels of skin pigmentation. Where relevant and available we present data by device type in Supporting Information Table S6, there may be some variation in the amount of mean bias based on device used but data are too limited to draw further conclusions.

### Association between ethnicity and pulse oximetry accuracy

Twenty-two of the 32 studies (4910 participants) described participants by ethnicity rather than level of skin pigmentation[31,33,34,36,38,39,41,45–50,55–63] and we included 14 studies (3510 participants) in meta-analyses.[31,33,34,39,45–47,49,50,59,60–63]. Pooled data are shown in Table 2 (further data are reported in Supporting Information Figures S5-S7 and Table S5).

Oxygen saturation measured for people described in study reports as Black or African American may be overestimated using hospital pulse oximetry compared with standard SaO_2_ readings: mean bias 1.52% (95% CI 0.95% to 2.09%), low-certainty evidence. The 95% confidence interval of this estimate ranges between an overestimation of 1% to 2%. The evidence for people described in studies as Asian, Hispanic, or of mixed ethnicity does not indicate a clinically important systematic bias (mean bias 0.31%, 0.09% to 0.54%), but it is of low certainty. The evidence is uncertain for groups described in papers as White/Caucasian, meaning further research is likely to alter findings (very low certainty evidence).

In the studies which referred to ethnicity rather than skin pigmentation, the *A*_*rms*_ values are around 2% or lower (95% CI non-estimable) for all groups, and the pooled SD values are around 1.50% on average (Table 2). This means that for people from the ethnic groups studied, about 68% of their pulse oximetry readings would be within ±2% of the CO-oximetry readings, with one SD indicating a variation around the mean bias of minus 1.50% to plus 1.50%.

We tested the sensitivity of the findings: A_*rms*_ and SD values were generally consistent but there was increased uncertainty for mean bias findings. Supporting Information Figure S8 presents evidence for each type of pulse oximeter evaluated: overall, most devices overestimated oxygen saturation in people described as Black or African American.

Eight of the 22 studies (1400 participants) were not included in meta-analysis (Supporting Information Table S7):[36,38,41,48,550-58] all had no accuracy data reported by ethnicity groups.

## Discussion

### Summary of findings

This review summarises the evidence from 32 studies which measure the impact of skin pigmentation (15 studies) or ethnicity (22 studies) on the accuracy of pulse oximetry. With mean bias as the primary outcome, the evidence suggests that for people with darkly pigmented skin and people described in study reports as Black or African American, oxygen saturation may be overestimated by pulse oximetry in hospital compared with gold standard SaO_2._ Whilst the existing evidence suggests that the degree of measurement bias (when rounded integers) is small at around the recommended criterion of 2% or less, this evidence is of low certainty and further confirmatory primary research is needed. It should also be emphasised that these results are for clinician-measured oximetry in controlled clinical environments and do not necessarily reflect the measurement bias of home pulse oximetry by patients or carers. The evidence suggests that pulse oximetry for people with other levels of skin pigmentation is less likely to be overestimated but the evidence is uncertain. The low certainty for much of the data presented means that further research could overturn these conclusions. For all the subgroups of populations evaluated, whilst the degree of mean bias is small or negligible, pulse oximetry readings appear unacceptably imprecise (pooled SDs are greater than the recommended criterion of 1%). Nevertheless, when the extents of measurement bias and precision are considered jointly in *A*_*rms*_, pulse oximetry measurements for all the subgroups appear acceptably accurate (with *A*_*rms*_ < the British Standards Institution recommended threshold of 4%).

### Overall completeness and applicability of evidence

Our search was comprehensive and up to date as of 14 December 2021 however, there are limitations in the completeness and applicability of the evidence identified.

Pulse oximetry is widely used in clinical practice and promoted for home use during the COVID-19 pandemic.[2] We are aware of many factors (such as oxygen saturation ranges at baseline, types of pulse oximeter probe, comorbidities, movement, age of the patient) which could theoretically affect pulse oximetry accuracy in the real world.[8] However, most of the included studies (27/32, 84.38%) in this review were based in hospital settings and this review only addresses skin pigmentation and ethnicity. Most included studies had limited information as to whether the pulse oximeters evaluated were appropriate for home self-monitoring. Therefore, little is known for the case of pulse oximetry undertaken by untrained people at home where other factors such as movement need to be considered.

Pulse oximeters have been developed and upgraded since 1970s,[65] and old devices might have been withdrawn. The included studies were published between 1985 and 2021 and some of the older studies may have used discontinued devices. Nevertheless, the overestimation of oxygen saturation for darker skin appears to be consistent in general across most devices evaluated. To keep the completeness of evidence in this review, we included study data for all pulse oximeter devices included.

We regarded mean bias as the primary outcome in this review given it is intuitive. We also present the results for the other outcomes recommended by British Standards Institution (overall accuracy and precision) for completeness.

### Comparison with other studies

To our knowledge, there is one existing systematic review (published in 1995) and a rapid review in this area.[6,8] The Jensen review used reference measures of SaO_2_ (PaO_2_, calculated SaO_2_, and %O_2_Hb) that are now considered incorrect or outdated.[12,15] The review considered skin pigmentation as a factor affecting pulse oximetry accuracy, but did not address ethnicity-related evidence. With no estimation of the size of the bias, the review concluded that pulse oximeters may overestimate oxygen saturation for people with darker skin. This conclusion however is only based on one study with data available for synthesis regarding skin pigmentation.

The rapid review by NHS Race and Health Observatory came to similar conclusions regarding pulse oximetry accuracy, but its methods were not clear. Of all nine studies included in the rapid review, two had no data on SpO_2_-SaO_2_ comparisons: one with a diagnostic accuracy design,[7] and one comparing two types of pulse oximetry with each other[66]. This rapid review did not differentiate between evidence for ethnicity and skin pigmentation, which is considered inappropriate.[14]

By comparison we identified 15 eligible studies exploring the influence of skin pigmentation and added the evidence of 22 studies which examined the influence of ethnicity. Importantly, we considered internationally recognised SaO_2_ measured by the gold standard CO-oximetry as the comparator for pulse oximetry. Our review, with more studies, adds well-produced evidence to the NHS Race and Health Observatory guidance and other national guidelines with further insights into the current size and precision around current accuracy estimates as noted below.

The UK British Standards Institution 2019, originating from the 2017 standards of the International Organization for Standardization (ISO),[67] specify performance requirements of pulse oximeter devices:[12] they are *A*_*rms*_ within 4% (the US Food and Drug Administration recommend a more conservative threshold of 3%)[68] and the mean bias (precision) value of

+/-2% (one SD) for acceptable pulse oximetry. In relation to these, the review results suggest an acceptable level of *A*_*rms*_ and bias but unacceptable imprecision. However, there is a need to consider the uncertainty in the mean bias estimates.

Whilst this is a relatively small amount of mean bias its impact on clinical decision making could be significant at threshold values for diagnosis of hypoxaemia, leading to clinically important hypoxaemia remaining undetected and untreated. Underestimated SpO_2_ readings also have the potential to be harmful, resulting in unnecessary treatment with oxygen (and the risk of hyperoxaemia) and wider impacts such as delayed hospital discharge. Two recent diagnostic studies provide evidence on clinical implications resulting from the bias in pulse oximetry for blood oxygen saturation levels[7,69]. In these studies people described as Black had a higher risk of ‘*occult hypoxemia that was not detected by pulse oximetry*’ compared with those described as White.[7] This may suggest that even small amounts of mean bias, when at the margins of diagnostic thresholds, could have an impact on diagnostic accuracy. Further understanding of these impacts could be explored via evidence synthesis of diagnostic accuracy (classification) studies to assess the clinical implications of the findings in relation to clinical decision-making thresholds.

The bias of pulse oximetry in specific sub-populations might have further implications. A recent decision analytical Markov modelling study evaluated the cost-utility of remote pulse oximetry monitoring over a 3-week time horizon from a US health sector perspective.[70] It found that remote monitoring dominates current standard care, by saving USD $11472 and gaining QALYs of 0.013. Besides, people with access to remote monitoring had 87% fewer hospitalisations and 77% fewer deaths. However, this study did not consider the pulse oximetry bias in specific sub-populations and future research is needed.

### Strengths and limitations of the review

We followed prespecified methods to summarise all relevant evidence to minimise the risk of bias in the review process.[11] We ran comprehensive electronic searches with the support of an information specialist, searched trial registries, and checked references of included studies and relevant systematic reviews identified. We used British Standards Institution recommended gold standard CO-oximetry as the comparator for pulse oximetry measures. We contacted study authors to clarify details and request study data. We developed a correlated hierarchical effects model and used RVE approaches to meta-analyse data.[21] This approach deals with correlations of multiple effect size estimates within a study, rather than treating them as independent (this treatment is common but inappropriate if conventional inverse variance analytical approach is chosen).[19] As a result of using RVE approaches, we included not only independent data (of 11 studies) in analyses but also more data from studies (n = 21) with repeated-measures design.

This review has some limitations. Firstly, seven studies considering the impact of skin pigmentation and eight studies considering ethnicity compared SpO_2_-SaO_2_ bias data between different subgroups and presented only tests of significance results, rather than SpO_2_ and SaO_2_ data per se at each subgroup level. Also, at least two studies used diagnostic accuracy design that only presented proportions of participants with specific ranges of SpO_2_ in relation to specific SaO_2_ values, again rather than SpO_2_ and SaO_2_ data per se [7,69]. We contacted authors of these studies to request relevant data and received data for two studies.[39,51] If more data were received, then the results of this review could change.

Secondly, we are aware of the potential difference between the concepts of race and ethnicity and the recent call to distinguish them from each other.[71] For simplicity, we chose to use the term of ‘ethnicity’ throughout this review given race and ethnicity are context/country-specific concepts and there is no globally accepted racial/ethnic classification approaches to clearly distinguish them.[14] If we had treated race and ethnicity data separately, the evidence base would change slightly, however, we would not expect the overall conclusion to change. We also acknowledge that measurement of level of skin pigmentation is limited by the use of scales like the Fitzpatrick scale.[72] Such scales are criticised as being too blunt a measure of skin pigmentation, an issue that impacts on the findings of this review and which needs to be considered in research going forward.

Thirdly, in meta-analysis, we grouped data from studies where people were grouped by ethnicity and referred to as ‘Asian’, ‘Hispanic’ or mixed ethnicity into a single subgroup; mainly because the included studies mixed these ethnic groups together in this way. We were not able to produce evidence for each of these ethnic groups separately, although we acknowledge potential differences in measurement bias between them.

Fourthly, we did not consider the differences between specific pulse oximeter devices (54 models included), the differences between children and adults and their health conditions, or the difference in skin pigmentation measurement methods between studies (Table 1 and Supporting Information Table S4). In terms of pulse oximeters evaluated, there may be differences between devices for the use of health professionals in hospitals and those for home self-monitoring. Because of these, meta-analyses in this review demonstrated between-studies heterogeneity inevitably (Table 2). However, we found, across devices evaluated and types of participants, included studies were largely consistent in suggesting oxygen saturation overestimation of using pulse oximetry. We therefore chose to pool study data, without undertaking further subgroups for these differences.

Fifthly, we only searched for English language publications. However, there are probably no major differences between summary treatment effects in English-language restricted meta-analyses and other language-inclusive meta-analyses.[73] We did not search for literature in the format of preprints or resources other than trial registries given the limited time and resources available. We considered the possible publication bias in assessing the certainty of evidence using GRADE approach.

Finally, there is no existing approach to risk of bias and GRADE assessment in the case of this review. We used the QUADAS-2 and GRADE approaches developed for the test accuracy topic: both are relevant but not specific to this review. However, we were only able to assess the certainty of evidence for mean bias and found the existing GRADE approach inapplicable for precision, *A*_*rms*_ and limits of agreement.

### Conclusions

Pulse oximetry may overestimate blood oxygen saturation levels for people with dark skin compared with gold standard SaO_2_ measures. The extent of measurement bias appears relatively small, but pulse oximetry measurements appear accurate for all levels of skin pigmentation when the small or negligible bias and imprecision identified are considered jointly in the overall accuracy.

The evidence for the measurement bias identified however is uncertain. A small bias may be crucial for some patients: particularly at the threshold that informs clinical decision making. Health professionals should consider the potential bias of pulse oximetry readings for people with dark skin, and should also be aware that multiple potential factors could affect the accuracy of pulse oximeter readings.

### Ethical approval

Not required.

### Data Availability

All relevant data are within the manuscript and its Supporting Information files. No additional data available.

## Acknowledgments

We thank the information specialist Catherine Harris for helping design search strategies and run electronic database searches in August 2021. We also thank Dr John Feiner and Dr Paul Young for sharing their study data and others who responded to our queries for clarifying study methods in deciding the eligibility of their studies.

## Author Contributions

**Conceptualization**: Chunhu Shi, Mark Goodall, Jo Dumville, Andrew Clegg, Caroline Leigh Watkins, George Georgiou, Catherine Elizabeth Lightbody, Nicky Cullum

**Data Curation**: Chunhu Shi

**Investigation**: Chunhu Shi, Mark Goodall, James Hill, Gill Norman, Oliver Hamer

**Formal analysis**: Chunhu Shi, Jo Dumville, Alexander Hodkinson

**Funding acquisition**: Jo Dumville, Caroline Leigh Watkins, Nicky Cullum

**Methodology**: Chunhu Shi, Jo Dumville, James Hill, Gill Norman, Andrew Clegg, Alexander Hodkinson

**Project administration**: Chunhu Shi, Jo Dumville, James Hill

**Resources**: Jo Dumville, Andrew Clegg, George Georgiou

**Software**: Chunhu Shi

**Supervision:** Jo Dumville, Andrew Clegg, Caroline Leigh Watkins, Nicky Cullum

**Validation**: Alexander Hodkinson, Jo Dumville, Gill Norman

**Visualization**: Chunhu Shi

**Writing – original draft**: Chunhu Shi, Jo Dumville, Nicky Cullum

**Writing – review & editing**: Chunhu Shi, Mark Goodall, Jo Dumville, James Hill, Gill Norman, Oliver Hamer, Andrew Clegg, Alexander Hodkinson, Caroline Leigh Watkins, George Georgiou, Catherine Elizabeth Lightbody, Paul Dark, Nicky Cullum

## Funding

The review is funded by the National Institute for Health Research Applied Research Collaboration (NIHR ARC) Greater Manchester and NIHR ARC North West Coast (ARC NWC). The views expressed in this article are those of the authors and not necessarily those of the National Institute for Health Research or the Department of Health and Social Care.

## Competing interests

The authors have declared that no competing interests exist.

## Notes

### Competing Interest Statement

The authors have declared no competing interest.

### Clinical Protocols

https://osf.io/gm7ty

