## Supporting Information for "The effects of skin pigmentation on the accuracy of pulse oximetry in measuring oxygen saturation: a systematic review and meta-analysis"

Table S1. Measure, definition, and data formats to assess the accuracy of pulse oximetry compared with reference measures

Box S1. The Ovid MEDLINE search strategy

Box S2. Data items in the data extraction form

Box S3. The QUADAS-2 tool used for assessing risk of bias and applicability (with further explanations in Notes)

Box S4. Data synthesis methods and generic R codes used

Table S2. Characteristics of the included studies

Table S3. Types of pulse oximeters and CO-oximetry evaluated in the included studies

Table S4. Mapping terms originally used for indicating skin pigmentation into low, medium or high level of skin pigmentation defined in the review for meta-analysis

Figure S1. Summary presentations of study sample sizes (n) and numbers of data pairs compared (N), accuracy root mean square ($A_{rms}$), mean bias (SD) and limits of agreement (LoA) of pulse oximeters for the subgroup of high (dark) skin pigmentation

Figure S2. Summary presentations of study sample sizes (n) and numbers of data pairs compared (N), accuracy root mean square ($A_{rms}$), mean bias (SD) and limits of agreement (LoA) of pulse oximeters for the subgroup of medium skin pigmentation

Figure S3. Summary presentations of study sample sizes (n) and numbers of data pairs compared (N), accuracy root mean square ($A_{rms}$), mean bias (SD) and limits of agreement (LoA) of pulse oximeters for the subgroup of low (light) skin pigmentation

Table S5. Summary of findings table for the impact of skin pigmentation and ethnicity on the accuracy of pulse oximetry compared with CO-oximetry

Figure S4. Summary presentations of study sample sizes (n) and numbers of data pairs compared (N), accuracy root mean square ($A_{rms}$), mean bias (SD) and limits of agreement (LoA) of pulse oximeters for levels of skin pigmentation by the different types of pulse oximeters

Table S6. Evidence from studies where skin pigmentation measures cannot be specified or grouped into low, medium, and/or high pigmentation

Figure S5. Summary presentations of study sample sizes (n) and numbers of data pairs compared (N), accuracy root mean square ($A_{rms}$), mean bias (SD) and limits of agreement (LoA) of pulse oximeters for the subgroup of Black/African American ethnic groups

Figure S6. Summary presentations of study sample sizes (n) and numbers of data pairs compared (N), accuracy root mean square ($A_{rms}$), mean bias (SD) and limits of agreement (LoA) of pulse oximeters for the subgroup of non-Black, non-White ethnic groups

Figure S7. Summary presentations of study sample sizes (n) and numbers of data pairs compared (N), accuracy root mean square ($A_{rms}$), mean bias (SD) and limits of agreement (LoA) of pulse oximeters for the subgroup of White/Caucasian ethnic groups

Figure S8. Summary presentations of study sample sizes (n) and numbers of data pairs compared (N), accuracy root mean square ($A_{rms}$), mean bias (SD) and limits of agreement (LoA) of pulse oximeters for ethnic groups by the different types of pulse oximeters

Table S7. Evidence from studies that could not be included in quantitative data pooling for the ethnicity factor

**Table S1. Measure, definition, and data formats to assess the accuracy of pulse oximetry compared with reference measures**

| Measure | Definition | Eligible data |
|---|---|---|
| The overall accuracy of pulse oximeter equipment | Agreement between a test result and an accepted reference value. That is a combination of a random component and of a common systematic error or bias component. | Expressed as the root-mean-square difference between measured values ($SpO_2$) and reference values ($SaO_2$). |
| Bias | Overestimation or underestimation of test measurement method relative to a reference measure. That is the total systematic error. | Assessed as mean difference between two measures (between-test mean, in this case $SaO_2$-$SpO_2$): a larger systematic difference means a larger bias value. |
| Precision | Closeness of agreement between independent test results obtained, which reflects the variability between test measures. Precision depends on random errors. | Reported as the standard deviation of between-test mean difference and a larger standard deviation means less precision. |
| Agreement | Concordance between two sets of measurements | Expressed as the limits of agreement (via the use of Bland-Altman plots) between different measures |

**Box S1. The Ovid MEDLINE search strategy**

1 exp Oximetry/
2 (oximet* or oxymet*).ti,ab,kw.
3 (SpO2 or %spo2 or sp o2).tw.
4 or/1-3
5 (co-oximet* or co-oxymet* or h?emoximet*).ti,ab,kw.
6 (blood adj3 (analys* or measure*)).tw.
7 (blood sampl* or gold standard or reference device* or reference instrument* or in-line oximet* or in vitro oximet* or arterial oxygen saturation or arterial oxyhemoglobin saturation or arterial oxyhaemoglobin saturation or arterial blood or arterial puncture or SaO2 or %SaO2 or sa o2).tw.
8 or/5-7
9 Reproducibility of Results/
10 Validation Study/
11 Evaluation Studies as Topic/
12 Bias/
13 "Sensitivity and Specificity"/
14 Hypoxia/di [Diagnosis]
15 comparative study.pt.
16 (accura* or inaccura* or overestimat* or over-estimat* or underestimat* or under-estimat* or agreement or root-mean-square or root mean square or RMS or quadratic mean).tw.
17 (precision or evaluat* or predict* or reliab* or reproducib* or concordance or performance or bias or validat* or error* or erroneous or individual variability or (variability and (analysis or values)) or sensitivity or specificity or failure).tw.
18 (compar* adj3 (measure* or value*)).tw.
19 (controlled desaturation or paired repeated measure* or method comparison or calibration stud*).ti,ab,kw.
20 (paired readings or paired measurements or "difference of values" or "limits of agreement" or "limits of values" or confidence limits or regression or bland altman).ti,ab,kw.
21 or/9-20
22 4 and 8 and 21
23 exp animals/ not humans.sh.
24 22 not 23
25 limit 24 to english language

**Box S2. Data items in the data extraction form**

- basic characteristics of studies, including first author, publication type, publication year, accuracy study type (lab-based, controlled desaturation study in healthy volunteers vs real-world accuracy study in patients);
- study setting;
- characteristics of participants, including eligibility criteria particularly health conditions, the number of participants and/or the number of pairs of oxygen saturation measures, and average age of participants;
- pulse oximetry tests being compared, including oximeters used and their manufacturers and models if available, probe type (transmissive vs reflectance probe), and location of sensor (e.g. finger, ear, toe);
- methods used for measuring $SaO_2$ including the blood gas analyser and CO-oximeter model, and the blood source such as the radial artery;
- skin pigmentation definitions including scales used and pigmentation levels, as well as, those for race/ethnicity;
- data on accuracy, bias and precision of measurement at a study level including comparative data based on level of skin pigmentation and that of race/ethnicity group as reported
- other outcome data available by level of skin pigmentation or by race/ethnicity groups
- unit of analysis (either individuals, or repeated measurements); and
- other factors that were reported to have the effects on pulse oximetry accuracy.

**Box S3. The QUADAS-2 tool used for assessing risk of bias and applicability (with further explanations in Notes)**

**Domain 1. Participant selection**: could the selection of participants have introduced bias?

Signalling questions for assessing risk of bias:

1. *Was an appropriate sample of participants included in the study?* [a]

2. Was a case-control design avoided? (omitted) [b]

3. Did the study avoid inappropriate exclusions? [c]

Signalling questions for assessing applicability:

- Are there concerns that the included participants and settings do not match the review question?

**Domain 2. Index test**: could the conduct or interpretation of the index test have introduced bias?

Signalling questions for assessing risk of bias:

4. *Were the pulse oximetry results interpreted without knowledge of the results of the reference standard?*

5. If a threshold was used, was it pre-specified? (omitted) [d]

Signalling questions for assessing applicability:

- Are there concerns that the index test, its conduct, or its interpretation differs from the review question?

**Domain 3. Reference standard**: could the reference standard, its conduct, or its interpretation have introduced bias?

Signalling questions for assessing risk of bias:

6. *Is the reference standard likely to correctly measure the blood oxygen saturation level?* [e]

7. *Were the reference standard results interpreted without knowledge of the results of pulse oximetry?*

Signalling questions for assessing applicability:

- Are there concerns that the target condition as defined by the reference standard does not match the question?

**Domain 4. Flow and timing**: could the analysis of flow and timing have introduced bias?
Signalling questions for assessing risk of bias:

8. *Was there an appropriate interval between pulse oximetry and reference standard? In this case it is considered appropriate that the index and reference standard measures are taken at the same time with no obvious time interval between them.*

9. Did all patients receive a reference standard?

10. Did all patients receive the same reference standard?

11. Were all patients included in the analysis? [f]

Notes:

a. To make the question more relevant for this accuracy review the wording has been amended. BSI for pulse oximetry (2019) allows for two types of participants for evaluating the SpO2 accuracy: healthy volunteers in a controlled desaturation study, and patients in clinical care settings. A study needs to define, select and recruit participants of interests accordingly. A study involving patients in clinical care settings may use consecutive or random sampling.

b. We omitted this question as it is more relevant to diagnostic test accuracy (DTA) reviews than this accuracy question.

c. For the pulse oximetry accuracy review, some studies may aim to evaluate the accuracy of pulse oximetry in people with a range of characteristics (e.g. level of skin pigmentation, baseline oxygen saturation levels), but inappropriately exclude important subgroups. This inappropriate exclusion may result in biased results in pulse oximetry accuracy. Therefore, new criteria may be applied to this signalling question even though the wording is unchanged from the original QUADAS-2 tool.

d. We omitted this question as it is more relevant for DTA reviews than this accuracy review.

e. Classification is particularly relevant to DTA. Pulse oximetry is used to measure (not classifying) SpO2. Therefore, the question is slightly re-worded by replacing ‘classify’ with ‘measure’.

For the pulse oximetry accuracy topic it is more relevant to consider how likely pulse oximetry SpO2 measurement follows recommended procedures, and if applicable, is carried out under appropriately standardised conditions. For example, BSI (2019) states, when the oxygen saturation needs to be changed to another level, there needs to be at least 30 s to allow SaO2 to reach stability before the pulse oximeter reading is taken. Similarly, blood sampling can begin only when the blood saturation stabilises at an acceptable level.

f. In this review we considered whether excluded data are considered ‘eligible’ for exclusion (with appropriate justifications) according to BSI for pulse oximetry guidance (2019). For example, for pulse oximeter monitors that set up an upper limit on displayed SpO2 (e.g. 99 %), data collected with SaO2 values beyond the specified SpO2 limit legitimately be excluded. Data pairs can be excluded if they were taken under conditions that were outside of the pre-planned test scope.

**Box S4. Data synthesis methods and generic R codes used**

**Part 1. Data synthesis methods used**

In this review, we performed meta-analyses for mean bias of $SpO_2$-$SaO_2$ and their SDs across included studies and used their pooled estimates to calculate overall accuracy and 95% limits of agreement. We described specific methods used below.
(1) Meta-analysis for mean bias of $SpO_2$-$SaO_2$ and SDs
We pooled study data of mean bias of $SpO_2$-$SaO_2$ for assessing whether use of pulse oximetry would under-estimate (pooled mean bias < 0) or over-estimate (pooled mean bias > 0) oxygen saturation in relation to CO-oximetry. We pooled study data on the SDs of mean bias for assessing the precision of pulse oximetry measures. For either mean bias or SD data pooling, we used the random-effects, correlated hierarchical effects model with small-sample corrections of the Robust Variance Estimation (RVE).[Pustejovsky 2021a] We chose this approach because

- we expected to include studies with repeated measures design, thus obtaining multiple dependent effect size estimates – data on mean bias and SD – within a study;
- we did not expect to have data on correlations between multiple dependent effect size estimates within a study as such correlations are commonly not reported in included studies. Therefore, the exact dependence structure of the multiple effect sizes within a study is unknown, and the conventionally used multivariate meta-analysis with known dependence structure of effect sizes could not be used in the case of this review.
- The approach we chose is a hybrid of correlated hierarchical effects model and RVE methods. Correlated hierarchical effects model allowed us to construct a flexible variance structure that could better capture two types of dependence: hierarchical effects and correlated effects. The RVE framework used modelling of dependence to approximate the unknown dependence structure and the structure does not have to be fully correct. Even if the structure is mis-specified, RVE's regression coefficient estimates (effect size) could be unbiased and standard errors could validly quantify uncertainty.[Hedges 2010] RVE approach uses products of the regression residuals to roughly approximate the variance-covariance structure of the errors (i.e. producing standard errors).[Pustejovsky 2021a] This estimation of standard errors is separated from the choice of weight matrices and therefore, standard errors can be produced without having to know the dependence structure.

More specifically we used the following methods to analyse data:
First, we used the method described by Tipton and Shuster to adjust dependent standard deviations, by which the under-estimation bias of the true standard deviation could be reduced in the case of repeated measures design.[Tipton 2017]
To obtain mean bias and its variance estimates for each study,

- mean bias = $SpO_2$-$SaO_2$ difference as reported
- $adjusted\ SD^2 = \ reported\ SD^2[(the\ total\ number\ of\ repeated\ measures - 1)/ (the\ total\ number\ of\ repeated\ measures - the\ number\ of\ replications\ per\ participant)]$

  E.g. in a study with 10 participants and a total of 200 repeated measures for a pulse oximetry, the number of replications per participant is 20 (i.e. 200/10).
- the sampling variance of mean bias = adjusted $SD^2$ / the number of participant.

In producing SD and its variance estimates for each study, we used their log-transform to normalise the distribution and stabilise its variance:

- log(adjusted $SD^2$) $\approx \log(adjusted\ SD^2) + \frac{1}{the\ numbe\ of\ participants - 1}$
- the sampling variance of log(adjusted $SD^2$) $\approx \frac{2}{the\ numbe\ of\ participants - 1}$

Second, we performed analyses in this section using rma.mv() function available in the package of *metafor.*[Viechtbauer 2010]

For pooling data on either mean bias or SD, we performed multi-level random-effects models without moderators for the dataset of either each level of skin pigmentation or ethnic group, where there was at least one study with at least two sets of effect size estimates (i.e. mean bias, or SD). For this, we used restricted maximum likelihood estimation in the function rma.mv().
By including a random term, multi-level random-effects model is specifically designed to deal with dependence among multiple effect size estimates within a study.[Viechtbauer 2010] We considered multiple effect size estimates within a study having random effects, thus including its indicator as a random term in each model to deal with their dependence.
For constructing reasonable multi-level random-effects models, we specified variance components with a correlation of 0.90. The correlation value resulted from the only systematic review in the same topic as our review.[Jensen 1995]
We used the $Tau^2$ statistic, produced by multi-level random-effects models, to quantify heterogeneity.

Third, based on multi-level random-effects model outputs, we finally used RVE approaches – more specifically the package of *clubSandwich*[Pustejovsky 2021b] – to estimate the RVE standard errors.

(2) Calculations of overall accuracy and 95% limits of agreement
There was no acceptable meta-analysis method to produce overall accuracy and 95% limits of agreement directly from study-level data of mean bias and SD for the case of this review.
When obtaining the pooled mean bias and the pooled SD for either each level of skin pigmentation or each ethnic group, we calculated overall accuracy using the BSI recommended method:[BSI 2019]

- $A_{rms}$ for the overall accuracy = $\sqrt{(the\ pooled\ mean\ bias^2\ +\ the\ pooled\ SD^2)}$

We also calculated the 95% limits of agreement using the following method described by Bland and Altman.[Bland 1986]

- 95% limits of agreement = pooled mean bias ± 1.96 * pooled SD

**Part 2. Generic R codes (bold and *Italic* texts) used for meta-analyses**

### Load R packages required
***library(clubSandwich)*** ### This package was for the robust variance estimation (RVE) approach
***library(metafor)***

### Load corresponding data used for meta-analysis of each level of skin pigmentation and each ethnicity group, respectively

dataset

### Run a multilevel random effects model (constant sampling correlation) for either bias or SD

```
V_mat <- impute_covariance_matrix(dataset$Vi,
                        cluster = dataset$level1,
                        r = 0.9,
```

```
                    smooth_vi = TRUE)

multilevel_model <- rma.mv(yi ~ 1,
                V = V_mat,
                random = ~ 1 | level1/ level2,
                data = dataset, sparse = TRUE, slab=paste(level1))

multilevel_model ### standard errors produced by these were model-based, rather than RVE ouputs

## the estimation CIs of tau2 for between-studies heterogeneity and within-study heterogeneity

confint(multilevel_model)
```

### the calculation of $I^2$ for an overall model

```
W <- diag(1/dataset$Vi)
X <- model.matrix(multilevel_model)
P <- W - W %*% X %*% solve(t(X) %*% W %*% X) %*% t(X) %*% W
100 * sum(multilevel_model$sigma2) / (sum(multilevel_model$sigma2) + (multilevel_model$k-multilevel_model$p)/sum(diag(P)))

## the separation of the overall I2 for between-studies and within-study heterogeneity

100 * multilevel_model$sigma2 / (sum(multilevel_model$sigma2) + (multilevel_model$k-multilevel_model$p)/sum(diag(P)))

## the calculation of RVE standard errors

CI_multilevel_model <- conf_int(multilevel_model, vcov = "CR2")
CI_multilevel_model
```

**Table S2. Characteristics of the included studies**

| Study | Study (test) settings | Pulse oximeter models evaluated | CO-oximetry | Blood source | Participant inclusion criteria | Average age | Factors of interest and measurements | Other factors |
|---|---|---|---|---|---|---|---|---|
| Abrams 2002 * | NR | 1 model:<br>Nellcor N-200 | Radiometer ABL520 | Arterial blood | Adult patients with cirrhosis (n = 294) | Mean 51.7 years | Race (White, Black) | Cirrhosis or not, oxygen saturation levels, hemoglobin levels, hepatopulmonary syndrome or not |
| Avant 1997 | Hospital or wards | 2 models:<br>Nellcor Oxiband,<br>Nellcor Dura-Y | CO-oximeter | Arterial blood | Critically ill children (n = 50) | Mean 26 months | Race (White, Black) | NR |
| Adler 1998 * | A & E | 1 model:<br>Nellcor D-25 | 4-wavelength spectro-photometer, or co-oximeter (Radiometer OSM3) | Arterial blood (no further detail) | Adult patients who needed blood gas analysis (n = 284) | Mean 60 years | Skin pigmentation measured using the Munsell colour system with categories of light, medium, or dark | NR |
| Bickler 2005 | Lab | 5 models:<br>Nellcor N-595 with Nellcor OxiMax A finger probe;<br>Two types of Novametrix 513s models;<br>Two types of Nonin Onyx models | Radiometer OSM3 | Arterial blood (radial or other arteries) | Healthy, non-smoking volunteers (n = 21) | Mean 29.05 years | Race/ethnicity and skin pigmentation with categories of light (northern European) and dark (African-American) | Oxygen saturation levels |

| | | | | | | | | |
|---|---|---|---|---|---|---|---|---|
| Bothma 1996 * | Hospital or wards | 3 models: Simed S100e; Nihon Koden; Ohmeda 3740 | IL482 co-oximeter | Arterial blood (no further detail) | Darkly pigmented critically ill adult patients (n = 100) | Adults, age not reported | All dark pigmentation objectively quantified using EEL reflectance spectrophotometer (Evans Electroselenium Company) | NA |
| Brooks 2020 | Hospital or wards | 2 models: Masimo, Nellcor (Covidien) | Radiometer ABL800 co-oximeter | Arterial blood | ICU infants and children (n = 929) | Median 1.9 years | Ethnicity (Aboriginal and/or Torres Strait Islander (ATSI), not ATSI) | Health conditions, age at admission, weight at admission, sex, sensor type (Massimo, Nellcor), SaO2 category, lactate, total haemoglobin (Hb), pH, oxygen saturation index, ventilation, inotropes, vasodilators, and vasoconstrictors |
| Ebmeier 2018 * | Hospital or wards | 2 models: Masimo oximeter for GE Marquette Rac-4A monitor; Philips sensors for Philips IntelliVue MP70 monitor | Radiometer ABL 800 FLEX arterial blood gas analyser | Arterial blood (no further detail) | Consecutive ICU patients (n = 394) | Mean 62.5 years | Skin pigmentation measured using the Fitzpatrick scale with categories of light (score of 1 or 2), medium (score of 3 or 4), and dark (score of 5 or 6) | $PaO_2$, acute physiology and chronic health evaluation (APACHE) II illness severity score, use of vasopressors, use of inotropes, capillary refill time (> vs < 3 seconds), body temperature, temperature of the hands, mean arterial |

| | | | | | | | | |
|---|---|---|---|---|---|---|---|---|
| | | | | | | | | pressure, pulse pressure, local factors |
| Escourrou 1990 | Hospital or wards | 3 models: Ohmeda Biox 3700; Criticare CSI 501+; Nellcor N-200 | Radiometer OSM2 | Arterial blood (radial or other arteries) | Adult patients with chronic pulmonary diseases (n = 101) | Range: 17 to 81 years | Skin pigmentation with categories of moderate vs unclear (but not Black) level | Exercise loads |
| Feiner 2007 | Lab | 3 models (6 types of finger probe): Nellcor N-595 (OxiMax A adhesive probe); Nellcor N-595 (a clip-type probe); Masimo Radical (clip probe); Masimo Radical (adhesive probe); Nonin 9700 (clip-type probe); Nonin 9700 (adhesive probe) | Radiometer OSM3 | Arterial blood (radial or other arteries) | Healthy non-smoking volunteers (n = 36) | Mean 29 years | Race/ethnicity and skin pigmentation defined as light (Caucasian), intermediate (Hispanic, Indian, Filipino, Vietnamese), and dark (African American) categories | Oxygen saturation levels, gender |
| Foglia 2017 * | Hospital or wards | 2 models: Nellcor Oximax (Covidien); Masimo Rainbow SET Radical 7 | Siemens Rapidlab 1265 | Arterial blood (no further detail) | Infants with cyanotic congenital heart disease and oxygen saturation <90% (n = 36) | Mean 6 days in light pigment, 118 days in dark pigment | Skin pigmentation measured using the Munsell Soil Book of Colour, Hue 7.5YR, with categories of light and dark | Oxygen saturation levels |
| Gabrielczyk 1988 | Hospital or wards | 1 model: Nellcor N-100 | Radiometer OSM2 | Arterial blood (radial or | Patients with postoperative hypothermia (n = 21) | Mean 59.5 years | Skin pigmentation with categories of racially pigmented | NR |

| | | | | | | | | |
|---|---|---|---|---|---|---|---|---|
| | | | | other arteries) | | | skin vs unclear pigmentation level | |
| Harris 2016 | Hospital or wards | 3 models: Masimo SET with LNCS sensor; Masimo SET Blue sensor; Nellcor N-600 Max-I sensor | AVOXimeter 1000E co-oximeter | Arterial blood (no further detail) | Hypoxemic pediatric patients with cyanotic congenital heart disease (n = 50) | Mean 18 months | Skin pigmentation measured using the Massey Skin Colour Score (categorised to be four levels) | Age, height, weight, binary indicators of non-White race and female gender |
| Harris 2019 * | Hospital or wards | 2 models: Masimo LNCS sensor; Nonin WristOx2 3150 with Bluetooth-enabled infant sensors 8008J | Bedside co-oximetry | Arterial blood (no further detail) | Hypoxemic infants with cyanotic heart disease (n = 24) | Median 13 days | Skin pigmentation measured using the Massey Skin Colour score (NR) | Age, sensor placement |
| Harskamp 2021 | Hospital or wards | 11 models: AFAC FS10D, AGPTEK FS10C, ANAPULSE ANP 100, Cocobear, Contec CMS50D1, HYLOGY MD-H37, Mommed YM101, PRCMISEMED F4PRO, PULOX PO-200, Zacurate Pro Series 500 DL, Philips M1191BL | Radiometer ABL90 Flex Plus | Arterial blood (radial or other arteries) | Intensive care patients (n = 35) | Mean 69 years | Skin pigmentation measured using the Fitzpatrick scale, with two categories: dark skin type (Fitzpatrick scale IV-VI) vs non-dark skin (Fitzpatrick I-III) | Age, sex, heart rate bias, body temperature, cold hands to touch, systolic blood pressure, and use of vasopressor drugs |
| Hinkelbein 2006 | Hospital or wards | 2 models: Nellcor DS-100A Durasensor sensor | Radiometer ABL625 | Arterial blood | ICU adults with mechanical | Mean 58.1 years | Race - all White (Caucasian) | NR |

| | | | | | | | | |
|---|---|---|---|---|---|---|---|---|
| | | with SIEMENS SC1281 monitor (SIREM module); Philips M1191A finger probe (PHILIPS IntelliVue MP70 monitor) | | | ventilation (n = 46) | | | |
| Hinkelbein 2007 * | Hospital or wards | 1 model: Nellcor DS-100A Durasensor sensor with SIEMENS SC1281 monitor (SIREM module) | Radiometer ABL625 | Arterial blood (radial or other arteries) | ICU adults with mechanical ventilation (n = 50) | Mean 59 years | Race - all White (Caucasian) | NR |
| Jubran 1990 | Hospital or wards | 2 models: Nellcor pulse oximeter, Ohmeda-Biox3700 pulse oximeter | CO-oximetry | Arterial blood | Critically ill, ventilator-dependent patients (n = 54) | Mean 53 years | Ethnicity – Black, and White categories | NR |
| Lee 1993 | Hospital or wards | 3 models: Nellcor, Simed, Critikon | Nova Stat Profile 3 pH/blood gas analyser | Arterial blood | ICU adults (n = 33) | Mean 56.4 years | Race (Chinese, Indian, Malay) | Hypoxia levels |
| McGovern 1996 | Hospital or wards | 1 model: Ohmeda 3700 | IL 482 Co-oximeter | Arterial blood (radial or other arteries) | Adults with stable condition with severe COPD (n = 8) | Mean 63.2 years | Race - all White | Exercise workload |
| Munoz 2008 * | Hospital or wards | 1 model: Minolta Pulsox-7 | IL 682 co-oximeter | Arterial blood (radial or other arteries) | Adults under assessment for long-term home oxygen therapy (n = 846) | Mean 68.4 years | Race – all Caucasian | Arterial oxygen tension and PaCO2, methods of measuring oxygen saturation (Oximeter vs co-oximeter ) |

| | | | | | | | | |
|---|---|---|---|---|---|---|---|---|
| Pilcher 2020 and Ploen 2016 (abstract) * | Hospital or wards | 14 models: Carescape B450 monitor with Nellcor probe; GE Dash 3000; Masimo Radical 7; Masimo SET Quartz (unspecified); Masimo SET Quartz Q400; Nonin 2120; Nonin 2140; Nonin Avant (unspecified); Nonin Avant 4000; Nonin Avant 9700; Nonin Lifesense Medair; Novametrix Model 512; Ohmeda Biox 3700E with a GE TruSignal or Nellcor probe; Philips Intellivue MP70 with a GE TruSignal Nellcor or Philips probe; Welch Allyn with a Nellcor probe | Radiometer ABL800 | Arterial blood (no further detail) | Hospitalised adult patients (n = 400) | Mean 64.2 years | Skin pigmentation measured using the modified Fitzpatrick scale with categories of light, medium, or dark | Care setting, probe location, chronic respiratory failure-related health conditions, current tobacco smoking status, diabetes mellitus |

| | | | | | | | | |
|---|---|---|---|---|---|---|---|---|
| Ries 1985 | Hospital or wards | 2 models: Ohmeda Biox IIA; Hewlett-Packard 47201A oximeter | IL282 co-oximeter | Arterial blood (radial or other arteries) | Pulmonary patients who underwent clinical exercise testing (n = 136) | Adults, age not reported | Skin pigmentation measured using a semi-quantitative scale of light to dark | Exercise loads, CoHgb, $SaO_2$ ranges |
| Ries 1989 | Hospital or wards | 2 models: Ohmeda Biox III; Hewlett-Packard 47201A oximeter | CO-oximeter | Arterial blood (radial or other arteries) | Pulmonary patients who underwent clinical exercise testing (n = 136) | Adults, age not reported | Skin pigmentation measured using the Munsell colour system with categories of very light, light, average, and moderately dark or very dark | |
| Ross 2014 | Hospital or wards | 3 models: Masimo pulse oximeters with Masimo LNCS probe, Nellcor pulse oximeters with Nellcor OxiMax probes, Masimo oximeters with Nellcor OxiMax probes | Radiometer ABL800 and Rapidlab 1265 (Siemens Healthcare), IL Gem 3000 | Arterial blood | ICU, mechanically ventilated children with cyanotic congenital heart disease (CCHD) or acute hypoxemic respiratory failure (n = 225) | Median 1 month for CCHD children, and 37 months for acute respiratory failure children | Race (African American, Hispanic, White, Asian, Other) | CCHD, prolonged capillary refill, having a SpO2 between 81% to 85%, 86% to 90%, or 91% to 95% (compared with a SpO2 of 96% to 97%), male gender, the combination of Masimo oximeter with a Nellcor sensor, mean airway pressure, hemoglobin, PICU site, temperature, fraction of inspired oxygen, age <2 months |

| | | | | | | | | |
|---|---|---|---|---|---|---|---|---|
| Schallom 2018 | Hospital or wards | 2 models: Nellcor OxiMax Forehead sensor, Xhale Assurance nasal alar sensor | Radiometer ABL800 Flex Series blood gas analyser | Arterial blood | Critically ill adults (n = 43) | Mean 60.1 years | Ethnicity | NR |
| Smyth 1986 | Lab | 2 models: Hewlett-Packard oximeter, Ohmeda Biox II oximeter | Corning 175- blood gas analyser co-oximeter | Arterial blood (radial or other arteries) | Healthy Caucasian volunteers (n = 6) | Range: 23 to 33 year | Race – all Caucasian | Oximeter model, oxygen saturation levels |
| Stewart 1991 | Hospital or wards | 1 model: Ohmeda Biox 3700 | Radiometer OSM2 | Arterial blood (radial or other arteries) | Adults with chronic rheumatic heart disease (n = 42) | Adults, age not reported | Ethnicity - all Chinese | Presence of triscupid regurgitation, pulse oximeter location |
| Thrush 1994 | Lab | 4 models: Critikon Dinamap Plus Model 8700, Critikon Oxyshuttle, Ohmeda 3700, MiniOx IV | IL482 co-oximeter | Arterial blood (radial or other arteries) | Healthy, non-smoking adults (n = 22) | Mean 29 years | Race – all White | Hypoxemia severity |
| Valbuena 2021 (retrospective design) | Hospital or wards | Not reported | Not reported, blood gas analysis | Arterial blood | Adult patients with respiratory failure or COVID-19 (n = 1562) | Not reported, > 18 years | Ethnicity – White, Black, Hispanic, and Asian categories | Not reported |
| Vesoulis 2021 (retrospective design) | Hospital or wards | 1 model: Nellcor MAX-N adhesive $SpO_2$ sensor (Covidien) (Philips IntelliVue | Radiometer ABL800 Flex | Arterial blood | Preterm infants at neonatal intensive care unit (n = 294) | Median 4 days | Ethnicity – White and Black categories | Not reported |

| | | | | | | | | |
|---|---|---|---|---|---|---|---|---|
| | | MP70 or MX800 monitor) | | | | | | |
| Wiles 2021 (retrospective design) | Hospital or wards | 1 model: Nellcor probes (GE Healthcare B1x5 M/P monitor) | RAPIDpoint 500 analyser (Siemens Healthcare GmbH) | Arterial blood | Adult patients with COVID-19 pneumonitis (n = 194) | Mean 62 years | Ethnicity – Asian, Black, White, and other categories | Not reported |
| Zeballos 1991 * | Lab | 2 models: Hewlett-Packard 47201A; Ohmeda Biox IIA | IL282 co-oximeter | Arterial blood (radial or other arteries) | Healthy, non-smoking volunteers (n = 33) | Mean 19 years | Race – all dark pigmentation (Black volunteers) | Exercise levels, sea levels |

Notes. * all studies used a prospective design.

**Table S3. Types of pulse oximeters and CO-oximetry evaluated in the included studies**

| Items | Summary statistics (n (%)) |
|---|---|
| Pulse oximeter manufacturers or brands (32 studies) | |
| AFAC | 1 study (3.12%), including one model:<br>• AFAC FS10D [Harskamp 2021] |
| AGPTEK | 1 study (3.12%), including one model:<br>• AGPTEK FS10C [Harskamp 2021] |
| ANAPULSE | 1 study (3.12%), including one model:<br>• ANAPULSE ANP 100 [Harskamp 2021] |
| CocoBear | 1 study (3.12%), including one model:<br>• Cocobear [Harskamp 2021] |
| Contec | 1 study (3.12%), including one model:<br>• Contec CMS50D1 [Harskamp 2021] |
| Criticare | 1 study (3.12%), including one model:<br>• Criticare CSI 501+ [Escourrou 1990] |
| Critikon | 2 studies (6.25%), including two specified models:<br>• Oxyshuttle [Thrush 1994]<br>• Dinamap Plus 8700 [Thrush 1994]<br>• Unspecified model [Lee 1993] |
| GE Healthcare | 1 study (3.12%), including two models:<br>• Carescape B450 monitor with Nellcor probe [Pilcher 2020]<br>• GE Dash 3000 [Pilcher 2020] |
| Hewlett-Packard | 4 studies (12.50%), including one specified model:<br>• Hewlett-Packard 47201A [Ries 1985; Ries 1989; Zeballos 1991]<br>• Unspecified model [Smyth 1986] |
| HYLOGY | 1 study (3.12%), including one specified model:<br>• HYLOGY MD-H37 [Harskamp 2021] |
| Masimo | 8 studies (25.00%), including at least five specified models:<br>• Radical 7 or Rainbow SET Radical 7 [Feiner 2007; Foglia 2017; Pilcher 2020]<br>• Masimo SET Quartz Q400 [Pilcher 2020]<br>• Masimo SET with LNCS sensor [Harris 2016; Harris 2019];<br>• Masimo SET Blue sensor [Harris 2016];<br>• Masimo SET Quartz (unspecified model) [Pilcher 2020] |

| | |
|---|---|
| | • Unspecified model [Brooks 2020; Ebmeier 2018; Ross 2014] |
| Mommed | 1 study (3.12%), including one specified model:<br>• Mommed YM101 [Harskamp 2021] |
| MiniOx | 1 study (3.12%), including one model:<br>• MiniOx IV [Thrush 1994] |
| Minolta | 1 study (3.12%), including one model:<br>• Pulsox-7 [Munoz 2008] |
| Nellcor | 18 studies (56.25%), including ten specified models:<br>• D-25 [Adler 1998]<br>• Dura-Y [Avant 1997]<br>• DS-100A Durasensor sensor (SIEMENS SC1281 monitor) [Hinkelbein 2006; Hinkelbein 2007]<br>• MAX-N sensor (Philips IntelliVue MP70 or MX800 monitor) [Vesoulis 2021]<br>• N-100 [Abrams 2002; Gabrielczyk 1988]<br>• N-200 [Escourrou 1990]<br>• N-595 [Bickler 2005; Feiner 2007]<br>• N-600 [Harris 2016]<br>• Oxiband [Avant 1997]<br>• OxiMax [Foglia 2017; Ross 2014; Schallom 2018]<br>• Unspecified model [Brooks 2020; Jubran 1990; Lee 1993; Ross 2014; Wiles 2021] |
| Nihon Koden | 1 study (3.12%), including one model:<br>• Nihon Koden [Bothma 1996] |
| Novametrix | 2 studies (6.25%), including two models:<br>• Novametrix 512 [Pilcher 2020]<br>• Novametrix 513s [Bickler 2005] |
| Nonin | 4 studies (12.50%), including seven specified models:<br>• Nonin 2120 [Pilcher 2020]<br>• Nonin 2140 [Pilcher 2020]<br>• Nonin Avant 4000 [Pilcher 2020]<br>• Nonin Avant 9700 [Feiner 2007; Pilcher 2020]<br>• Nonin Lifesense Medair [Pilcher 2020]<br>• 3150 WristOx2 [Harris 2019]<br>• Onyx [Bickler 2005] |

| | |
|---|---|
| | • Nonin Avant (unspecified) [Pilcher 2020] |
| Ohmeda | 11 studies (34.38%), including six specified models:<br>• Biox II [Smyth 1986]<br>• Biox IIA [Ries 1985; Zeballos 1991]<br>• Biox III [Ries 1989]<br>• Biox 3700 [Escourrou 1990; Jubran 1990; McGovern 1996; Stewart 1991; Thrush 1994]<br>• Biox 3700E; [Pilcher 2020]<br>• 3740 [Bothma 1996] |
| Philips | 4 studies (12.50%), including two specified models:<br>• Philips M1191A (Philips IntelliVue MP70 monitor) [Hinkelbein 2006]<br>• Philips M1191BL [Harskamp 2021]<br>• Unspecified Philips oximeter (Philips IntelliVue MP70 monitor) [Ebmeier 2018; Pilcher 2020] |
| PRCMISEMED | 1 study (3.12%), including one model:<br>• PRCMISEMED F4PRO [Harskamp 2021] |
| PULOX | 1 study (3.12%), including one model:<br>• PULOX PO-200 [Harskamp 2021] |
| Simed | 2 studies (6.25%), including one specified model:<br>• S100e [Bothma 1996]<br>• Unspecified model [Lee 1993] |
| Welch Allyn | 1 study (3.12%), including one model:<br>• Welch Allyn [Pilcher 2020] |
| Xhale Assurance | 1 study (3.12%), including one model:<br>• Xhale Assurance [Schallom 2018] |
| Zacurate | 1 study (3.12%), including one model:<br>• Zacurate Pro Series 500 DL [Harskamp 2021] |
| Comparator co-oximeter devices (32 studies) | |
| AVOXimeter | 1 study (3.12%), including one method:<br>• AVOXimeter 1000E co-oximeter [Harris 2016] |
| Corning co-oximeter | 1 study (3.12%), including one method:<br>• Corning 175 blood gas analyser co-oximeter [Smyth 1986] |

| | |
|---|---|
| IL co-oximeter | 6 studies (18.75%), including three methods:<br>• IL 282 co-oximeter [Ries 1985; Zeballos 1991]<br>• IL 482 co-oximeter [Bothma 1996; McGovern 1996; Thrush 1994]<br>• IL 682 co-oximeter [Munoz 2008] |
| Nova Stat Profile co-oximeter | 1 study (3.12%), including one method:<br>• Nova Stat Profile 3 pH/blood gas analyser [Lee 1993] |
| Radiometer co-oximeter | 14 studies (43.75%), including six methods:<br>• Radiometer ABL520 [Abrams 2002]<br>• Radiometer ABL625 [Hinkelbein 2006; Hinkelbein 2007]<br>• Radiometer ABL800 Flex co-oximeter [Brooks 2020; Ebmeier 2018; Pilcher 2020; Schallom 2018; Vesoulis 2021]<br>• Radiometer ABL90 Flex Plus [Harskamp 2021]<br>• Radiometer OSM2 [Escourrou 1990; Gabrielczyk 1988; Stewart 1991]<br>• Radiometer OSM3 [Bickler 2005; Feiner 2007] |
| Siemens co-oximeter | 2 studies (6.25%), including two methods:<br>• Siemens Rapidlab 1265 analyser [Foglia 2017]<br>• Siemens RAPIDpoint 500 analyser [Wiles 2021] |
| Combinations of different co-oximeters | 2 studies (6.25%), including two methods:<br>• 4-wavelength spectro-photometer, or co-oximeter (Radiometer OSM3) [Adler 1998]<br>• Radiometer ABL800 and Rapidlab 1265 (Siemens Healthcare), IL Gem 3000 [Ross 2014] |
| Unspecified co-oximeter | 5 studies (15.62%), including:<br>• Unspecified methods [Avant 1997; Harris 2019; Jubran 1990; Ries 1989; Valbuena 2021] |

**Table S4. Mapping terms originally used for indicating skin pigmentation into low, medium or high level of skin pigmentation defined in the review for meta-analysis**

| Skin pigmentation measurement methods | The number of classification categories as reported | Low (light) skin pigmentation | Medium skin pigmentation | High (dark) skin pigmentation |
|---|---|---|---|---|
| Fitzpatrick scale [Ebmeier 2018; Pilcher 2020; Ploen 2016] | Three categories | 'Light (Type I to Type II)', or 'light (score of 1 or 2)' | 'Medium (Type III to Type IV)', or 'medium (score of 3 or 4)' | 'Dark (Type V to Type VI)', or 'dark (score of 5 or 6)' |
| Munsell colour system [Adler 1998; Foglia 2017] | Two categories [Foglia 2017] | 'Light' | - | 'Dark' |
| | Three categories [Adler 1998] | 'Light' | 'Medium' | 'Dark' |
| Using ethnicity to indicate skin pigmentation [Bickler 2005; Feiner 2007; Zeballos 1991] | One category [Zeballos 1991] | - | - | 'Black' participants |
| | Two categories [Bickler 2005] | 'Light (northern European)' | - | 'Dark (African-American)' |
| | Three categories [Feiner 2007] | 'Light (Caucasian)' | 'Intermediate (Hispanic, Indian, Filipino, Vietnamese)' | 'Dark (African American)' |
| Objective quantification using a reflectance spectrophotometer [Bothma 1996] | One category [Bothma 1996] | | | 'Dark pigmentation' |

**Figure S1. Summary presentations of study sample sizes (n) and numbers of data pairs compared (N), accuracy root mean square ($A_{rms}$), mean bias (SD) and limits of agreement (LoA) of pulse oximeters for the subgroup of high (dark) skin pigmentation**

| Study | Pulse oximeters | Sample (data pairs) | Arms, % | Mean bias (SD), % | LoA, % | Mean bias (95% CI), % |
|---|---|---|---|---|---|---|
| Ebmeier 2018 | Philips (IntelliVue MP70) | 8 (8) | 2.76 | 2.24 (1.73) | -1.15 to 5.63 | 2.24 (1.04 to 3.44) |
| Ebmeier 2018 | Masimo (GE Marquette Rac-4A) | 4 (4) | 1.15 | 1.10 (0.39) | 0.33 to 1.87 | 1.10 (0.72 to 1.48) |
| Foglia 2017 | Masimo Radical (Rainbow) | 14 (14) | 5.06 | 1.60 (4.80) | -7.81 to 11.01 | 1.60 (-0.99 to 4.19) |
| Foglia 2017 | Nellcor Oximax | 14 (14) | 7.43 | 5.40 (5.10) | -4.60 to 15.40 | 5.40 (2.81 to 7.99) |
| Feiner 2007 | Masimo Radical (clip) | 17 (372) | 3.63 | 2.14 (2.93) | -3.60 to 7.88 | 2.14 (1.00 to 3.28) |
| Feiner 2007 | Nellcor N-595 (clip) | 17 (371) | 2.91 | 2.04 (2.08) | -2.04 to 6.12 | 2.04 (0.90 to 3.18) |
| Feiner 2007 | Nonin 9700 (clip) | 17 (372) | 1.33 | -0.63 (1.17) | -2.92 to 1.66 | -0.63 (-1.77 to 0.51) |
| Feiner 2007 | Masimo Radical (adhesive) | 17 (372) | 2.81 | -0.24 (2.80) | -5.73 to 5.25 | -0.24 (-1.38 to 0.90) |
| Feiner 2007 | Nellcor N-595 (OxiMax, adhesive) | 17 (359) | 3.22 | 2.13 (2.42) | -2.61 to 6.87 | 2.13 (0.99 to 3.27) |
| Feiner 2007 | Nonin 9700 (adhesive) | 17 (371) | 2.45 | 1.26 (2.10) | -2.86 to 5.38 | 1.26 (0.12 to 2.40) |
| Bickler 2005 | Nonin Onyx | 7 (119) | 1.49 | 0.15 (1.48) | -2.75 to 3.05 | 0.15 (-1.24 to 1.54) |
| Bickler 2005 | Novametrix 513 | 7 (119) | 2.80 | 1.83 (2.12) | -2.33 to 5.99 | 1.83 (0.44 to 3.22) |
| Bickler 2005 | Nellcor N-595 | 11 (209) | 2.41 | 1.29 (2.03) | -2.69 to 5.27 | 1.29 (-0.10 to 2.68) |
| Adler 1998 | Nellcor D-25 | 34 (34) | 4.30 | -2.20 (3.70) | -9.45 to 5.05 | -2.20 (-3.44 to -0.96) |
| Bothma 1996 | Simed S100e | 100 (100) | 2.25 | 1.20 (1.90) | -2.52 to 4.92 | 1.20 (0.77 to 1.63) |
| Bothma 1996 | Nihon Koden | 100 (100) | 2.62 | 0.85 (2.48) | -4.00 to 5.70 | 0.85 (0.42 to 1.28) |
| Bothma 1996 | Ohmeda 3740 (finger) | 100 (100) | 2.05 | 0.55 (1.98) | -3.32 to 4.42 | 0.55 (0.12 to 0.98) |
| Bothma 1996 | Ohmeda 3740 (ear) | 100 (100) | 2.60 | -1.00 (2.40) | -5.70 to 3.70 | -1.00 (-1.43 to -0.57) |
| Zeballos 1991* | Hewlett-Packard 47201A | 33 (33) | 1.75 | -0.40 (1.70) | -3.73 to 2.93 | -0.40 (-2.28 to 1.48) |
| Zeballos 1991* | Ohmeda Biox IIA | 33 (33) | 2.90 | 2.10 (2.00) | -1.82 to 6.02 | 2.10 (0.22 to 3.98) |
| Zeballos 1991* | Hewlett-Packard 47201A | 11 (11) | 1.88 | -0.80 (1.70) | -4.13 to 2.53 | -0.80 (-2.68 to 1.08) |
| Zeballos 1991* | Ohmeda Biox IIA | 11 (11) | 4.48 | 3.50 (2.80) | -1.99 to 8.99 | 3.50 (1.62 to 5.38) |
| Zeballos 1991* | Hewlett-Packard 47201A | 22 (22) | 8.41 | -4.80 (6.90) | -18.32 to 8.72 | -4.80 (-6.68 to -2.92) |
| Zeballos 1991* | Ohmeda Biox IIA | 22 (22) | 11.99 | 9.80 (6.90) | -3.72 to 23.32 | 9.80 (7.92 to 11.68) |
| Primary analysis | 8 studies (24 comparisons) | 221 (3270) | 1.88 | 1.11 (1.52) | -1.87 to 4.09 | 1.11 (0.29 to 1.93) |
| Sensitivity (RoB) | 6 studies (15 comparisons) | 177 (2691) | 1.75 | 0.87 (1.52) | -2.11 to 3.84 | 0.87 (-0.46 to 2.19) |
| Sensitivity (Scale) | 5 studies (9 comparisons) | 160 (474) | 1.79 | 0.89 (1.55) | -2.16 to 3.93 | 0.89 (-1.37 to 3.14) |
| Sensitivity (Homo) | 6 studies (14 comparisons) | 88 (2738) | 1.95 | 1.12 (1.60) | -2.02 to 4.26 | 1.12 (-0.16 to 2.39) |

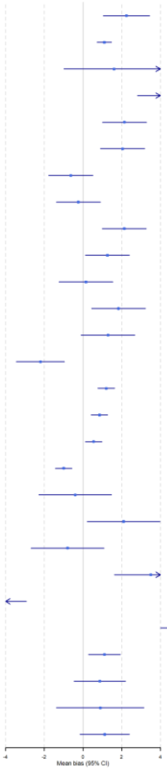

Note:

- The $Chi^2$ test for heterogeneity in the primary analysis suggested a Q(df = 23) = 2251.16, with P value < 0.0001.
- $Tau^2$ between the 8 studies = 0 (95% CI 0 to 3.77); $Tau^2$ between the 24 comparisons = 7.16 (4.14 to 13.81).
- The estimated overall $I^2$ for the primary analysis = 98.03%, of which about 0% is due to between-studies heterogeneity, and 98.03% due to within-study heterogeneity.

**Figure S2. Summary presentations of study sample sizes (n) and numbers of data pairs compared (N), accuracy root mean square ($A_{rms}$), mean bias (SD) and limits of agreement (LoA) of pulse oximeters for the subgroup of medium skin pigmentation**

| Study | Pulse oximeters | Sample (data pairs) | Arms, % | Mean bias (SD), % | LoA, % | Mean bias (95% CI), % |
|---|---|---|---|---|---|---|
| Pilcher 2020 | 14 models | 157 (157) | 1.94 | -1.10 (1.60) | -4.24 to 2.04 | -1.10 (-1.35 to -0.85) |
| Ebmeier 2018 | Philips (IntelliVue MP70) | 89 (89) | 2.15 | 0.45 (2.12) | -3.71 to 4.60 | 0.45 (0.01 to 0.89) |
| Ebmeier 2018 | Masimo (GE Marquette Rac-4A) | 48 (48) | 1.76 | 0.11 (1.77) | -3.37 to 3.59 | 0.11 (-0.39 to 0.61) |
| Feiner 2007 | Masimo Radical (clip) | 7 (154) | 2.32 | 0.99 (2.10) | -3.13 to 5.11 | 0.99 (-0.70 to 2.68) |
| Feiner 2007 | Nellcor N-595 (clip) | 7 (154) | 1.71 | 0.64 (1.59) | -2.48 to 3.76 | 0.64 (-1.05 to 2.33) |
| Feiner 2007 | Nonin 9700 (clip) | 7 (154) | 1.92 | 0.03 (1.92) | -3.73 to 3.79 | 0.03 (-1.66 to 1.72) |
| Feiner 2007 | Masimo Radical (adhesive) | 7 (154) | 1.99 | -0.34 (1.96) | -4.18 to 3.50 | -0.34 (-2.03 to 1.35) |
| Feiner 2007 | Nellcor N-595 & OxiMax (adhesive) | 7 (154) | 2.48 | 0.83 (2.34) | -3.76 to 5.42 | 0.83 (-0.86 to 2.52) |
| Feiner 2007 | Nonin 9700 (adhesive) | 7 (154) | 2.69 | 0.51 (2.64) | -4.66 to 5.68 | 0.51 (-1.18 to 2.20) |
| Adler 1998 | Nellcor D-25 | 105 (105) | 5.91 | -2.80 (5.20) | -12.99 to 7.39 | -2.80 (-3.79 to -1.81) |
| Primary analysis | 4 studies (10 comparisons) | 406 (1323) | 1.58 | -0.58 (1.47) | -3.46 to 2.30 | -0.58 (-2.25 to 1.09) |
| Sensitivity (Scale) | 3 studies (4 comparisons) | 399 (399) | 1.66 | -0.79 (1.46) | -3.66 to 2.08 | -0.79 (-3.03 to 1.45) |

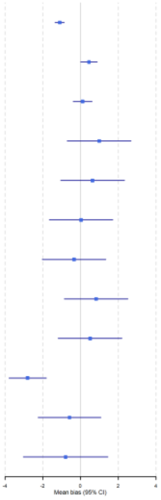

Note:

- The $Chi^2$ test for heterogeneity in the primary analysis suggested a Q(df = 9) = 81.92, with P value < 0.0001.
- $Tau^2$ between the 4 studies = 1.47 (95% CI 0 to 10.91); $Tau^2$ between the 10 comparisons = 0.18 (0.02 to 1.17).
- The estimated overall $I^2$ for the primary analysis = 92.65%, of which about 82.39% is due to between-studies heterogeneity, and 10.25% due to within-study heterogeneity.

**Figure S3. Summary presentations of study sample sizes (n) and numbers of data pairs compared (N), accuracy root mean square ($A_{rms}$), mean bias (SD) and limits of agreement (LoA) of pulse oximeters for the subgroup of low (light) skin pigmentation**

| Study | Pulse oximeters | Sample (data pairs) | Arms, % | Mean bias (SD), % | LoA, % | Mean bias (95% CI), % |
|---|---|---|---|---|---|---|
| Pilcher 2020 | 14 models | 242 (242) | 2.08 | -1.20 (1.70) | -4.53 to 2.13 | -1.20 (-1.41 to -0.99) |
| Ebmeier 2018 | Philips (IntelliVue MP70) | 98 (98) | 2.17 | 0.17 (2.19) | -4.12 to 4.45 | 0.17 (-0.27 to 0.60) |
| Ebmeier 2018 | Masimo (GE Marquette Rac-4A) | 142 (142) | 2.19 | -1.07 (1.92) | -4.83 to 2.69 | -1.07 (-1.39 to -0.75) |
| Foglia 2017 | Masimo Radical (Rainbow) | 21 (21) | 3.81 | 0.20 (3.80) | -7.25 to 7.65 | 0.20 (-1.76 to 2.16) |
| Foglia 2017 | Nellcor OxiMax | 19 (19) | 5.83 | 3.00 (5.00) | -6.80 to 12.80 | 3.00 (1.04 to 4.96) |
| Feiner 2007 | Masimo Radical (clip) | 12 (263) | 1.73 | 0.21 (1.72) | -3.16 to 3.58 | 0.21 (-0.79 to 1.21) |
| Feiner 2007 | Nellcor N-595 (clip) | 12 (263) | 1.71 | 0.36 (1.67) | -2.91 to 3.63 | 0.36 (-0.64 to 1.36) |
| Feiner 2007 | Nonin 9700 (clip) | 12 (263) | 1.78 | -0.95 (1.51) | -3.91 to 2.01 | -0.95 (-1.95 to 0.05) |
| Feiner 2007 | Masimo Radical (adhesive) | 12 (263) | 2.34 | -0.64 (2.25) | -5.05 to 3.77 | -0.64 (-1.64 to 0.36) |
| Feiner 2007 | Nellcor N-595 & OxiMax (adhesive) | 12 (263) | 1.87 | 1.04 (1.55) | -2.00 to 4.08 | 1.04 (0.04 to 2.04) |
| Feiner 2007 | Nonin 9700 (adhesive) | 12 (263) | 1.45 | 0.52 (1.35) | -2.13 to 3.17 | 0.52 (-0.48 to 1.52) |
| Bickler 2005 | Nonin Onyx | 9 (200) | 1.44 | -0.21 (1.42) | -2.99 to 2.57 | -0.21 (-1.40 to 0.98) |
| Bickler 2005 | Novametrix 513 | 9 (200) | 2.13 | 1.15 (1.79) | -2.36 to 4.66 | 1.15 (-0.04 to 2.34) |
| Bickler 2005 | Nellcor N-595 | 10 (220) | 2.07 | -0.27 (2.05) | -4.29 to 3.75 | -0.27 (-1.46 to 0.92) |
| Adler 1998 | Nellcor D-25 | 145 (145) | 5.24 | -2.50 (4.60) | -11.52 to 6.52 | -2.50 (-3.25 to -1.75) |
| Primary analysis | 6 studies (15 comparisons) | 670 (2865) | 1.53 | -0.35 (1.49) | -3.27 to 2.58 | -0.35 (-1.36 to 0.67) |
| Sensitivity (RoB) | 5 studies (12 comparisons) | 660 (2245) | 1.57 | -0.47 (1.50) | -3.42 to 2.47 | -0.47 (-1.77 to 0.83) |
| Sensitivity (Scale) | 4 studies (6 comparisons) | 648 (667) | 1.62 | -0.54 (1.53) | -3.53 to 2.45 | -0.54 (-2.52 to 1.43) |

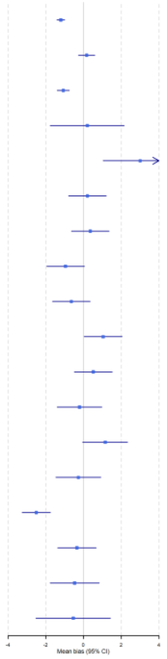

Note:

- The $Chi^2$ test for heterogeneity in the primary analysis suggested a Q(df = 14) = 243.72, with P value < 0.0001.
- $Tau^2$ between the 6 studies = 0.32 (95% CI 0 to 4.87); $Tau^2$ between the 15 comparisons = 0.99 (0.37 to 2.96).
- The estimated overall $I^2$ for the primary analysis = 92.73%, of which about 22.42% is due to between-studies heterogeneity, and 70.31% due to within-study heterogeneity.

**Table S5. Summary of findings table for the impact of skin pigmentation and ethnicity on the accuracy of pulse oximetry compared with CO-oximetry**

| Outcomes | **Anticipated absolute effects*** (95% CI) | | Mean bias (mean $SpO_2$-$SaO_2$) (95% CI) | № of participants (studies) | Certainty of the evidence (GRADE) | Comments |
|---|---|---|---|---|---|---|
| | **Oxygen saturation measured by pulse oximetry ($SpO_2$)** | **Actual oxygen saturation measured by CO-oximetry ($SaO_2$)** | | | | |
| Mean bias in people with high (dark) skin pigmentation | 90% | **89%** (90% to 88%) | **Mean bias 1.11** (0.29 to 1.93) | 221 participants with 3270 $SpO_2$-$SaO_2$ pairs (8 studies with 24 comparison evaluations) | ⊕⊕⊕⊖ Moderate [a] | Pulse oximetry $SpO_2$ readings probably overestimate arterial oxygen saturation by on average 1.11% compared with the $SaO_2$ measure of CO-oximetry in people with high (dark) skin pigmentation. |
| Mean bias in people with medium skin pigmentation | 90% | **90%** (89% to 92%) | **Mean bias -0.58** (-2.25 to 1.09) | 406 participants with 1323 $SpO_2$-$SaO_2$ pairs (4 studies with 10 comparison evaluations) | ⊕⊖⊖⊖ Very low [a,b] | It is uncertain if pulse oximetry would overestimate arterial oxygen saturation compared with the use of CO-oximetry in people with medium skin pigmentation. |
| Mean bias in people with low (light) skin pigmentation | 90% | **90%** (89% to 91%) | **Mean bias -0.35** (-1.36 to 0.67) | 670 participants with 2865 $SpO_2$-$SaO_2$ pairs (6 studies with 15 comparison evaluations) | ⊕⊕⊖⊖ Low[a,c] | Pulse oximetry may not overestimate arterial oxygen saturation compared with the use of CO-oximetry in people with low (light) skin pigmentation. |
| Mean bias in people from Black/African American ethnic groups | 90% | **89%** (89% to 88%) | **Mean bias 1.52** (0.95 to 2.09) | 459 participants with 5753 $SpO_2$-$SaO_2$ pairs (9 studies with 22 comparison evaluations) | ⊕⊕⊖⊖ Low [d,e] | Pulse oximetry $SpO_2$ readings may overestimate arterial oxygen saturation by on average 1.52% compared with the $SaO_2$ measure of CO-oximetry in people from Black/African American ethnic groups. |
| Mean bias in people of ethnicity other than Black or White such as Asians, Hispanics, those of mixed ethnicity | 90% | **90%** (90% to 89%) | **Mean bias 0.31** (0.09 to 0.54) | 522 participants with 2646 $SpO_2$-$SaO_2$ pairs (3 studies with 9 comparison evaluations) | ⊕⊕⊖⊖ Low [d,e] | Pulse oximetry may very slightly overestimate arterial oxygen saturation compared with the use of CO-oximetry in people of ethnicity other than Black and White such as Asians, Hispanics, those of mixed ethnicity. |

| Outcomes | Anticipated absolute effects* (95% CI) | | Mean bias (mean $SpO_2$-$SaO_2$) (95% CI) | № of participants (studies) | Certainty of the evidence (GRADE) | Comments |
|---|---|---|---|---|---|---|
| | **Oxygen saturation measured by pulse oximetry ($SpO_2$)** | **Actual oxygen saturation measured by CO-oximetry ($SaO_2$)** | | | | |
| Mean bias in people from White/Caucasian ethnic groups | 90% | **89%** (89% to 90%) | **Mean bias 0.55** (-0.21 to 1.31) | 2195 participants with 12870 $SpO_2$-$SaO_2$ pairs (13 studies with 48 comparison evaluations) | ⊕⊖⊖⊖ Very low[a,e,f] | It is uncertain if pulse oximetry would overestimate or underestimate arterial oxygen saturation compared with the use of CO-oximetry in White/Caucasians. |

***The actual oxygen saturation measured by CO-oximetry ($SaO_2$)** (and its 95% confidence interval) is based on the assumed oxygen saturation measured by pulse oximetry ($SpO_2$) of 90% and the **relative effect** (and its 95% CI).

**GRADE Working Group grades of evidence**
**High certainty:** we are very confident that the true effect lies close to that of the estimate of the effect.
**Moderate certainty:** we are moderately confident in the effect estimate: the true effect is likely to be close to the estimate of the effect, but there is a possibility that it is substantially different.
**Low certainty:** our confidence in the effect estimate is limited: the true effect may be substantially different from the estimate of the effect.
**Very low certainty:** we have very little confidence in the effect estimate: the true effect is likely to be substantially different from the estimate of effect.

### *Explanations*

a. Downgraded once for the joint consideration of inconsistency and publication bias. Firstly, the analysis found either high statistical heterogeneity, differences between studies in pulse oximetry devices, and/or the large variation of point estimates on the forest plot. Secondly, despite a comprehensive search, only part of the included studies presented data for meta-analysis and only English-language publications were searched for.

b. Downgraded twice for imprecision. The limits of the CI are very large and cover values that lead to different conclusions on pulse oximetry's accuracy: e.g., the lower limit suggests a clear underestimation whilst the upper limit suggests a clear overestimation.

c. Downgraded once for imprecision. The limits of the CI are slightly wide and the range covers values that potentially lead to different conclusions on pulse oximetry's accuracy: e.g., the lower limit suggests a small underestimation. The upper limit suggests a small overestimation.

d. Downgraded once for the joint consideration of study limitations and publication bias. Firstly, a proportion of the included studies and data were at high overall risk of bias. Secondly, despite a comprehensive search, only part of the included studies presented data for meta-analysis and only English-language publications were searched for.

e. Downgraded once for the indirectness. The evidence from data synthesis for ethnic groups was indirectly relevant to the topic of skin pigmentation for this review.

f. Downgraded once for study limitations. In the meta-analysis, around half of the included studies and/or data were at high overall risk of bias.

**Figure S4. Summary presentations of study sample sizes (n) and numbers of data pairs compared (N), accuracy root mean square ($A_{rms}$), mean bias (SD) and limits of agreement (LoA) of pulse oximeters for levels of skin pigmentation by the different types of pulse oximeters**

| | Studies (comparisons) | Sample (data pairs) | Arms, % | Mean bias (SD), % | LoA, % | Mean bias (95% CI), % |
|---|---|---|---|---|---|---|
| * Masimo | | | | | | |
| Low pigmentation | 3 studies (4 comparisons) | 175 (689) | 1.57 | -0.57 (1.47) | -3.44 to 2.31 | -0.57 (-2.66 to 1.53) |
| Medium pigmentation | 2 studies (3 comparisons) | 55 (356) | 1.34 | 0.19 (1.32) | -2.40 to 2.78 | 0.19 (-1.14 to 1.52) |
| High pigmentation | 3 studies (4 comparisons) | 35 (762) | 1.93 | 1.12 (1.57) | -1.96 to 4.20 | 1.12 (0.43 to 1.81) |
| * Nellcor | | | | | | |
| Low pigmentation | 4 studies (5 comparisons) | 186 (910) | 1.80 | 0.09 (1.79) | -5.22 to 5.40 | 0.09 (-3.40 to 3.58) |
| Medium pigmentation | 2 studies (3 comparisons) | 112 (413) | 2.24 | -1.09 (1.95) | -7.17 to 4.99 | -1.09 (-23.50 to 21.40) |
| High pigmentation | 4 studies (5 comparisons) | 76 (987) | 2.29 | 1.51 (1.72) | -5.07 to 8.09 | 1.51 (-3.26 to 6.27) |
| * Nonin | | | | | | |
| Low pigmentation | 2 studies (3 comparisons) | 21 (726) | 1.27 | -0.21 (1.25) | -2.67 to 2.25 | -0.21 (-0.24 to -0.18) |
| Medium pigmentation | 1 study (2 comparisons) | 7 (308) | 1.62 | 0.27 (1.60) | -2.86 to 3.40 | 0.27 (-1.54 to 2.08) |
| High pigmentation | 2 studies (3 comparisons) | 24 (862) | 1.31 | 0.26 (1.29) | -2.26 to 2.78 | 0.26 (-0.73 to 1.25) |
| * Philips | | | | | | |
| Low pigmentation | 1 study (1 comparison) | 98 (98) | 2.17 | 0.17 (2.19) | -4.12 to 4.45 | 0.17 (-0.27 to 0.60) |
| Medium pigmentation | 1 study (1 comparison) | 89 (89) | 2.15 | 0.45 (2.12) | -3.71 to 4.60 | 0.45 (0.01 to 0.89) |
| High pigmentation | 1 study (1 comparison) | 8 (8) | 2.76 | 2.24 (1.73) | -1.15 to 5.63 | 2.24 (0.80 to 3.68) |
| * Novametrix | | | | | | |
| Low pigmentation | 1 study (1 comparison) | 9 (200) | 2.13 | 1.15 (1.79) | -2.36 to 4.66 | 1.15 (0.90 to 1.40) |
| High pigmentation | 1 study (1 comparison) | 7 (119) | 2.80 | 1.83 (2.12) | -2.33 to 5.99 | 1.83 (1.45 to 2.21) |
| * Ohmeda | | | | | | |
| High pigmentation | 2 studies (5 comparisons) | 133 (266) | 2.98 | 2.55 (1.53) | -4.08 to 9.18 | 2.55 (-31.50 to 36.60) |
| * Nihon | | | | | | |
| High pigmentation | 1 study (1 comparison) | 100 (100) | 2.62 | 0.85 (2.48) | -4.00 to 5.70 | 0.85 (0.37 to 1.34) |
| * Simed | | | | | | |
| High pigmentation | 1 study (1 comparison) | 100 (100) | 2.25 | 1.20 (1.90) | -2.52 to 4.92 | 1.20 (0.83 to 1.57) |
| * Hewlett-Packard | | | | | | |
| High pigmentation | 2 studies (23 comparisons) | 33 (66) | 2.55 | -2.00 (1.59) | -5.11 to 1.11 | -2.00 (-5.24 to 1.24) |
| * A group of 14 models | | | | | | |
| Low pigmentation | 1 study (1 comparison) | 242 (242) | 1.56 | -1.20 (1.70) | -4.53 to 2.13 | -1.20 (-1.41 to -0.99) |
| Medium pigmentation | 1 study (1 comparison) | 157 (157) | 1.49 | -1.10 (1.60) | -4.24 to 2.04 | -1.10 (-1.35 to -0.85) |
| High pigmentation | 1 study (1 comparison) | 1 (1) | NA | 0.10 (NA) | NA | 0.10 (NA) |

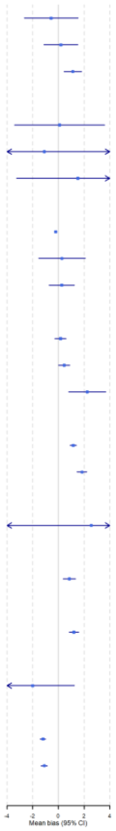

This figure presents the impact of skin pigmentation on pulse oximetry accuracy according to types of pulse oximeters evaluated. Results of analyses suggested that:

- Masimo, Nellcor, Philips, Novametrix, Ohmeda, Nihon, and Simed appear to have higher $SpO_2$ readings than $SaO_2$ by on average 1% in people with high skin pigmentation.
- Hewlett-Packard appears to produce $SpO_2$ measures 2% lower than $SaO_2$ readings in people with high skin pigmentation.
- Novametrix produces a $SpO_2$ measure higher than $SaO_2$ readings by on average 1% in people with low (light) skin pigmentation whilst others of these devices appear to produce $SpO_2$ measures with a bias no more than 1% compared with $SaO_2$ readings in people from medium and low skin pigmentation subgroups.
- Nonin does not result in over- or underestimation of oxygen saturation in people with any level of skin pigmentation.

### Table S6. Evidence from studies where skin pigmentation measures cannot be specified or grouped into low, medium, and/or high pigmentation

| Measures of skin pigmentation | No. of participants (data pairs) and no. of studies and evaluations | Summary of reported results | Comments |
|---|---|---|---|
| ***Subjective skin pigmentation categories related to ethnic groups ('moderately pigmented' or 'racially pigmented' skin, as reported, versus an unclear level of skin pigmentation)*** | 122 (267) in two studies with two evaluations [Escourrou 1990, Gabrielczyk 1988] | Skin pigmentation levels did not affect pulse oximetry accuracy:<br>• Across Ohmeda Biox 3700, Criticare CSI 501+, and Nellcor N200, the mean difference in bias between moderate pigmentation and others = 1.1% (SD 3), and skin pigmentation levels did not affect pulse oximetry accuracy.[Escourrou 1990]<br>• Nellcor N100 over-estimated oxygen saturation with a mean difference of 0.6%, and skin pigmentation levels did not affect pulse oximetry accuracy [ Gabrielczyk 1988] | **Four models evaluated*:*** Ohmeda Biox 3700, Criticare CSI 501+, Nellcor N200, Nellcor N100 |
| ***Massey score without specified categories*** | 74 (603) in two studies with five evaluations [Harris 2016; Harris 2019] | Of the four models evaluated,<br>• There were no significant findings for the Masimo Standard sensor, Nellcor N600, and the WristOx2 3150 sensor regarding the effect of skin pigmentation on pulse oximetry accuracy.[Harris 2016, Harris 2019]<br>• Multivariable regression models for the mean bias from the Masimo Blue sensor yielded a significant effect for a Massey Score of 4 ($p = 0.006$), indicating an increase in average bias of 4% relative to individuals with a score of 1, adjusting for other demographic factors.[Harris 2016] | **Four models evaluated:** Masimo SET LNCS sensor (Masimo Standard); Masimo SET Blue sensor; Nellcor N-600 with Max-I sensor; Nonin (Bluetooth-enabled) WristOx2 3150 with 8008J sensors |
| ***An unnamed 4-level system with original categories of light to dark that were grouped into moderate and light pigmentation by the study authors*** | 23 (198) in one study with four evaluations [Ries 1985] | Skin pigmentation was associated with the pulse oximetry accuracy for Hewlett-Packard ear oximeter 47201A but not for Biox IIA (Ohmeda)<br>• Differences between Hewlett-Packard oximeter readings and $SaO_2$ were significantly larger in the five darker pigmented participants (16 data pairs) than in the 18 light pigmented ones (83 data pairs, $t = -2.18$, $p < 0.05$). It is unclear whether the larger difference suggested overestimation or underestimation.<br>• $SpO_2$ (Biox IIA)-$SaO_2$ differences in darker pigmented participants were not significantly different from those in light pigmentation participants ($t = 1.68$, $p > 0.05$). | **Two models evaluated:** Hewlett-Packard 47201A ear oximeter;<br>Ohmeda Biox IIA |
| ***Munsell system with four categories that could not be classified into low, medium, and high level of pigmentation*** | 154 (973) in one study with eight evaluations [Ries 1989] | There was a higher overestimation of $SpO_2$ in using Ohmeda Biox III in people with high pigmentation than those with low pigmentation. However, Hewlett-Packard 47201A oximeter did not overestimate $SpO_2$ in either 'very light', 'light', 'average', or 'moderately dark or very dark' pigmentation groups of the Munsell system.<br>• Hewlett-Packard 47201A:<br>Very light – Arms = 3.23, mean bias (SD) = -0.30 (SD 3.22), limit of agreement = -6.61 to 6.01;<br>Light – Arms = 2.02, mean bias (SD) = 0 (2.02), limits of agreement = -3.96 to 3.96;<br>Average – Arms = 1.99, mean bias (SD) = -0.60 (1.90), limits of agreement = -4.32 to 3.12;<br>Moderately dark or very dark – Arms = 1.82, mean bias (SD) = -0.90 (1.58), limits of agreement = -4.00 to 2.20<br>• Ohmeda Biox III:<br>Very light – Arms = 2.40, mean bias (SD) = 0.60 (SD 2.32), limit of agreement = -3.95 to 5.15;<br>Light – Arms = 2.30, mean bias (SD) = 0.40 (2.27), limits of agreement = -4.04 to 4.84;<br>Average – Arms = 2.58, mean bias (SD) = 1.40 (2.17), limits of agreement = -2.85 to 5.65;<br>Moderately dark or very dark – Arms = 2.35, mean bias (SD) = 1.20 (2.02), limits of agreement = -2.76 to 5.16 | **Two models evaluated:** Hewlett-Packard 47201A oximeter;<br>Ohmeda Biox III<br><br>This study did not reported numbers of participants for each level of skin pigmentation |
| ***Fitzpatrick scale with four categories that were*** | 35 (2492) in one study with 11 | $SpO_2$-$SaO_2$ bias, a continuous measure, was used for a multivariable logistic regression including Fitzpatrick scale and analyses showed that, compared with Fitzpatrick scale I-III categories, Fitzpatrick IV-VI categories as a group was | |

| | | |
|---|---|---|
| ***classified by the authors to be two groups: IV-VI categories and I-III categories*** | evaluations [Harskamp 2021] | significantly associated with bias for five models: AGPTEK FS10C regression beta 1.96 (SE 0.93), p = 0.04; Cocobear beta 2.30 (1.21), p = 0.05; HYLOGY MD-H37 beta 3.07 (1.13), p = 0.007; Mommed YM101 beta 2.40 (0.82), p = 0.004; and Zacurate Pro Series 500DL beta 2.34 (1.1), p = 0.038. All beta values are positive and larger than 1, suggesting a higher skin pigmentation means a higher bias. However, the association was not significant for the other pulse oximetry models: AFAC FS10D, ANAPULSE ANP 100, Contec CMS50D1, PRCMISEMED F4 PRO, and PULOX-PO-200, all with p > 0.05 |

**Figure S5. Summary presentations of study sample sizes (n) and numbers of data pairs compared (N), accuracy root mean square ($A_{rms}$), mean bias (SD) and limits of agreement (LoA) of pulse oximeters for the subgroup of Black/African American ethnic groups**

| Study | Pulse oximeters | Sample (data pairs) | Arms, % | Mean bias (SD), % | LoA, % | Mean bias (95% CI), % |
|---|---|---|---|---|---|---|
| Valbuena 2021 | Unspecified | 195 (195) | 10.40 | 1.70 (10.20) | -18.70 to 22.20 | 1.70 (0.27 to 3.13) |
| Vesoulis 2021 | Nellcor MAX-N | 124 (2044) | 9.50 | 1.73 (9.34) | -16.58 to 20.04 | 1.73 (0.08 to 3.38) |
| Wiles 2021 | Nellcor (GE B1x5 M/P) | 19 (599) | 1.56 | 0.75 (1.39) | -3.47 to 1.97 | 0.75 (0.11 to 1.39) |
| Feiner 2007 | Masimo Radical (clip) | 17 (372) | 3.63 | 2.14 (2.93) | -3.60 to 7.88 | 2.14 (1.00 to 3.28) |
| Feiner 2007 | Nellcor N-595 (clip) | 17 (371) | 2.91 | 2.04 (2.08) | -2.04 to 6.12 | 2.04 (0.90 to 3.18) |
| Feiner 2007 | Nonin 9700 (clip) | 17 (372) | 1.33 | -0.63 (1.17) | -2.92 to 1.66 | -0.63 (-1.77 to 0.51) |
| Feiner 2007 | Masimo Radical (adhesive) | 17 (372) | 2.81 | -0.24 (2.80) | -5.73 to 5.25 | -0.24 (-1.38 to 0.90) |
| Feiner 2007 | Nellcor N-595 (OxiMax, adhesive) | 17 (359) | 3.22 | 2.13 (2.42) | -2.61 to 6.87 | 2.13 (0.99 to 3.27) |
| Feiner 2007 | Nonin 9700 (adhesive) | 17 (371) | 2.45 | 1.26 (2.10) | -2.86 to 5.38 | 1.26 (0.12 to 2.40) |
| Bickler 2005 | Nonin Onyx | 7 (119) | 1.49 | 0.15 (1.48) | -2.75 to 3.05 | 0.15 (-1.24 to 1.54) |
| Bickler 2005 | Novametrix 513 | 7 (119) | 2.80 | 1.83 (2.12) | -2.33 to 5.99 | 1.83 (0.44 to 3.22) |
| Bickler 2005 | Nellcor N-595 | 11 (209) | 2.41 | 1.29 (2.03) | -2.69 to 5.27 | 1.29 (-0.10 to 2.68) |
| Abrams 2002 | Nellcor N-200 | 16 (16) | 3.83 | 3.60 (1.30) | 1.05 to 6.15 | 3.60 (2.96 to 4.24) |
| Avant 1997 | Nellcor Oxiband | 15 (30) | 2.79 | 1.35 (2.44) | -3.43 to 6.13 | 1.35 (0.05 to 2.65) |
| Avant 1997 | Nellcor Dura-Y | 15 (30) | 2.79 | 1.05 (2.59) | -4.03 to 6.13 | 1.05 (-0.25 to 2.35) |
| Zeballos 1991* | Hewlett-Packard 47201A | 33 (33) | 1.75 | -0.40 (1.70) | -3.73 to 2.93 | -0.40 (-2.28 to 1.48) |
| Zeballos 1991* | Ohmeda Biox IIA | 33 (33) | 2.90 | 2.10 (2.00) | -1.82 to 6.02 | 2.10 (0.22 to 3.98) |
| Zeballos 1991* | Hewlett-Packard 47201A | 11 (11) | 1.88 | -0.80 (1.70) | -4.13 to 2.53 | -0.80 (-2.68 to 1.08) |
| Zeballos 1991* | Ohmeda Biox IIA | 11 (11) | 4.48 | 3.50 (2.80) | -1.99 to 8.99 | 3.50 (1.62 to 5.38) |
| Zeballos 1991* | Hewlett-Packard 47201A | 22 (22) | 8.41 | -4.80 (6.90) | -18.32 to 8.72 | -4.80 (-6.68 to -2.92) |
| Zeballos 1991* | Ohmeda Biox IIA | 22 (22) | 11.99 | 9.80 (6.90) | -3.72 to 23.32 | 9.80 (7.92 to 11.68) |
| Jubran 1990 | Nellcor/Ohmeda Biox 3700 | 29 (43) | 4.26 | 3.30 (2.70) | -2.00 to 8.60 | 3.30 (2.31 to 4.29) |
| Primary analysis | 9 studies (22 comparisons) | 459 (5753) | 2.27 | 1.52 (1.68) | -1.78 to 4.82 | 1.52 (0.95 to 2.09) |
| Sensitivity (RoB) | 4 studies (10 comparisons) | 67 (2892) | 1.99 | 1.47 (1.35) | -1.17 to 4.11 | 1.47 (-0.21 to 3.16) |
| Sensitivity (Trans) | 7 studies (20 comparisons) | 316 (3110) | 2.26 | 1.55 (1.64) | -1.67 to 4.77 | 1.55 (0.85 to 2.25) |
| Sensitivity (Homo) | 8 studies (16 comparisons) | 426 (5621) | 2.35 | 1.63 (1.69) | -1.68 to 4.94 | 1.63 (0.77 to 2.49) |

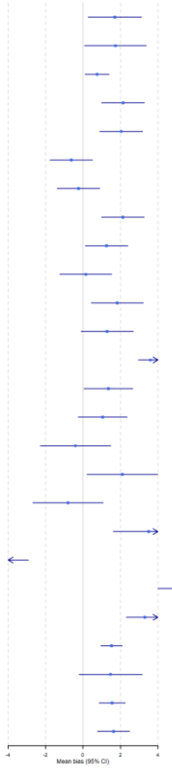

Note:

- The $Chi^2$ test for heterogeneity in the primary analysis suggested a Q(df = 21) = 1640.85, with P value < 0.0001.
- $Tau^2$ between the 9 studies = 0 (95% CI 0 to 3.04); $Tau^2$ between the 22 comparisons = 6.86 (3.91 to 13.58).
- The estimated overall $I^2$ for the primary analysis = 96.39%, of which about 0% is due to between-studies heterogeneity, and 96.39% due to within-study heterogeneity.

**Figure S6. Summary presentations of study sample sizes (n) and numbers of data pairs compared (N), accuracy root mean square ($A_{rms}$), mean bias (SD) and limits of agreement (LoA) of pulse oximeters for the subgroup of non-Black, non-White ethnic groups**

| Study | Pulse oximeters | Sample (data pairs) | Arms, % | Mean bias (SD), % | LoA, % | Mean bias (95% CI), % |
|---|---|---|---|---|---|---|
| Valbuena 2021 | Unspecified | 243 (243) | 5.80 | -0.30 (5.80) | -11.80 to 11.30 | -0.30 (-1.07 to 0.47) |
| Valbuena 2021 | Unspecified | 238 (238) | 6.50 | 0.80 (6.40) | -12.10 to 13.70 | 0.80 (0.03 to 1.57) |
| Wiles 2021 | Nellcor (GE B1x5 M/P) | 34 (1241) | 1.13 | 0.33 (1.09) | -2.47 to 1.80 | 0.33 (-0.04 to 0.70) |
| Feiner 2007 | Masimo Radical (clip) | 7 (154) | 2.32 | 0.99 (2.10) | -3.13 to 5.11 | 0.99 (-0.70 to 2.68) |
| Feiner 2007 | Nellcor N-595 (clip) | 7 (154) | 1.71 | 0.64 (1.59) | -2.48 to 3.76 | 0.64 (-1.05 to 2.33) |
| Feiner 2007 | Nonin 9700 (clip) | 7 (154) | 1.92 | 0.03 (1.92) | -3.73 to 3.79 | 0.03 (-1.66 to 1.72) |
| Feiner 2007 | Masimo Radical (adhesive) | 7 (154) | 1.99 | -0.34 (1.96) | -4.18 to 3.50 | -0.34 (-2.03 to 1.35) |
| Feiner 2007 | Nellcor N-595 (OxiMax, adhesive) | 7 (154) | 2.48 | 0.83 (2.34) | -3.76 to 5.42 | 0.83 (-0.86 to 2.52) |
| Feiner 2007 | Nonin 9700 (adhesive) | 7 (154) | 2.69 | 0.51 (2.64) | -4.66 to 5.68 | 0.51 (-1.18 to 2.20) |
| Primary analysis | 3 studies (9 comparisons) | 522 (2646) | 1.58 | 0.31 (1.55) | -2.72 to 3.35 | 0.31 (0.09 to 0.54) |
| Sensitivity (RoB) | 2 studies (7 comparisons) | 41 (2165) | 1.23 | 0.36 (1.18) | -1.95 to 2.66 | 0.36 (-0.24 to 0.95) |
| Sensitivity (Trans) | 2 studies (8 comparisons) | 488 (1405) | 2.07 | 0.30 (2.04) | -3.70 to 4.31 | 0.30 (-0.80 to 1.40) |

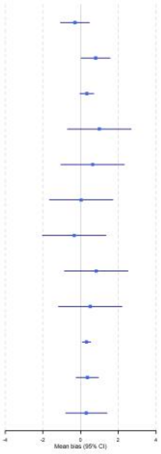

Note:

- The $Chi^2$ test for heterogeneity in the primary analysis suggested a Q(df = 8) = 56.20, with P value < 0.0001.
- $Tau^2$ between the 3 studies = 0 (95% CI 0 to 2.34); $Tau^2$ between the 9 comparisons = 0.23 (0.07 to 0.90).
- The estimated overall $I^2$ for the primary analysis = 47.95%, of which about 0% is due to between-studies heterogeneity, and 47.95% due to within-study heterogeneity.

**Figure S7. Summary presentations of study sample sizes (n) and numbers of data pairs compared (N), accuracy root mean square ($A_{rms}$), mean bias (SD) and limits of agreement (LoA) of pulse oximeters for the subgroup of White/Caucasian ethnic groups**

| Study | Pulse oximeters | Sample (data pairs) | Arms, % | Mean bias (SD), % | LoA, % | Mean bias (95% CI), % |
|---|---|---|---|---|---|---|
| Valbuena 2021 | Unspecified | 675 (675) | 7.30 | 0.30 (7.30) | -14.20 to 14.90 | 0.30 (-0.25 to 0.85) |
| Vesoulis 2021 | Nellcor MAX-N | 170 (2342) | 8.90 | 0.72 (8.87) | -16.67 to 18.11 | 0.72 (-0.62 to 2.06) |
| Wiles 2021 | Nellcor (GE B1x5 M/P) | 135 (4197) | 1.08 | -0.28 (1.06) | -1.79 to -2.35 | -0.28 (-0.46 to -0.10) |
| Munoz 2008 | Minolta Pulsox-7 | 846 (846) | 2.38 | 0.47 (2.33) | -4.10 to 5.04 | 0.47 (0.31 to 0.63) |
| Feiner 2007 | Masimo Radical (clip) | 12 (263) | 1.73 | 0.21 (1.72) | -3.16 to 3.58 | 0.21 (-0.79 to 1.21) |
| Feiner 2007 | Nellcor N-595 (clip) | 12 (263) | 1.71 | 0.36 (1.67) | -2.91 to 3.63 | 0.36 (-0.64 to 1.36) |
| Feiner 2007 | Nonin 9700 (clip) | 12 (263) | 1.78 | -0.95 (1.51) | -3.91 to 2.01 | -0.95 (-1.95 to 0.05) |
| Feiner 2007 | Masimo Radical (adhesive) | 12 (263) | 2.34 | -0.64 (2.25) | -5.05 to 3.77 | -0.64 (-1.64 to 0.36) |
| Feiner 2007 | Nellcor N-595 (OxiMax, adhesive) | 12 (263) | 1.87 | 1.04 (1.55) | -2.00 to 4.08 | 1.04 (0.04 to 2.04) |
| Feiner 2007 | Nonin 9700 (adhesive) | 12 (263) | 1.45 | 0.52 (1.35) | -2.13 to 3.17 | 0.52 (-0.48 to 1.52) |
| Hinkelbein 2007* | Nellcor DS-100A Durasensor | 50 (50) | 1.51 | -0.20 (1.50) | -3.14 to 2.74 | -0.20 (-0.87 to 0.47) |
| Hinkelbein 2007* | Nellcor DS-100A Durasensor | 50 (50) | 3.40 | -1.60 (3.00) | -7.48 to -4.28 | -1.60 (-2.27 to -0.93) |
| Hinkelbein 2007* | Nellcor DS-100A Durasensor | 50 (50) | 2.86 | -1.20 (2.60) | -6.30 to -3.90 | -1.20 (-1.87 to -0.53) |
| Hinkelbein 2007* | Nellcor DS-100A Durasensor | 50 (50) | 3.67 | -1.10 (3.50) | -7.96 to -5.76 | -1.10 (-1.77 to -0.43) |
| Hinkelbein 2007* | Nellcor DS-100A Durasensor | 50 (50) | 2.38 | -0.60 (2.30) | -5.11 to 3.91 | -0.60 (-1.27 to 0.07) |
| Hinkelbein 2007* | Nellcor DS-100A Durasensor | 50 (50) | 2.38 | -0.90 (2.20) | -5.21 to -3.41 | -0.90 (-1.57 to -0.23) |
| Hinkelbein 2007* | Nellcor DS-100A Durasensor | 50 (50) | 2.47 | -0.90 (2.30) | -5.41 to -3.61 | -0.90 (-1.57 to -0.23) |
| Hinkelbein 2007* | Nellcor DS-100A Durasensor | 50 (50) | 2.25 | -0.80 (2.10) | -4.92 to -3.32 | -0.80 (-1.47 to -0.13) |
| Hinkelbein 2007* | Nellcor DS-100A Durasensor | 50 (50) | 2.15 | -0.80 (2.00) | -4.72 to -3.12 | -0.80 (-1.47 to -0.13) |
| Hinkelbein 2007* | Nellcor DS-100A Durasensor | 50 (50) | 2.31 | -0.70 (2.20) | -5.01 to -3.61 | -0.70 (-1.37 to -0.03) |
| Hinkelbein 2006† | Nellcor DS-100A Durasensor | 23 (69) | 2.61 | -0.70 (2.51) | -5.62 to 4.22 | -0.70 (-1.89 to 0.49) |
| Hinkelbein 2006† | Philips M1191A | 23 (69) | 2.83 | 0.60 (2.77) | -4.83 to 6.03 | 0.60 (-0.59 to 1.79) |
| Hinkelbein 2006† | Nellcor DS-100A Durasensor | 23 (69) | 3.33 | -1.10 (3.14) | -7.25 to 5.05 | -1.10 (-2.29 to 0.09) |
| Hinkelbein 2006† | Philips M1191A | 23 (69) | 3.14 | 0.80 (3.04) | -5.16 to 6.76 | 0.80 (-0.39 to 1.99) |
| Bickler 2005 | Nonin Onyx | 9 (200) | 1.44 | -0.21 (1.42) | -2.99 to 2.57 | -0.21 (-1.40 to 0.98) |
| Bickler 2005 | Novametrix 513 | 9 (200) | 2.13 | 1.15 (1.79) | -2.36 to 4.66 | 1.15 (-0.04 to 2.34) |
| Bickler 2005 | Nellcor N-595 | 10 (220) | 2.07 | -0.27 (2.05) | -4.29 to 3.75 | -0.27 (-1.46 to 0.92) |
| Abrams 2002 | Nellcor N-200 | 184 (184) | 3.85 | 3.40 (1.80) | -0.13 to 6.93 | 3.40 (3.14 to 3.66) |
| Avant 1997 | Nellcor Oxiband | 35 (70) | 3.06 | 1.45 (2.69) | -3.82 to 6.72 | 1.45 (0.57 to 2.33) |
| Avant 1997 | Nellcor Dura-Y | 35 (70) | 2.87 | 1.21 (2.60) | -3.89 to 6.31 | 1.21 (0.33 to 2.09) |
| McGovern 1996 | Ohmeda 3700 | 8 (49) | 3.36 | -1.70 (2.90) | -7.38 to 3.98 | -1.70 (-3.83 to 0.43) |
| Thrush 1994‡ | Critikon Dinamap Plus 8700 | 22 (88) | 3.00 | 0.00 (3.00) | -5.88 to 5.88 | 0.00 (-1.15 to 1.15) |
| Thrush 1994‡ | Critikon Dinamap Plus 8700 | 22 (88) | 2.24 | 1.00 (2.00) | -2.92 to 4.92 | 1.00 (-0.15 to 2.15) |
| Thrush 1994‡ | Critikon Dinamap Plus 8700 | 22 (88) | 2.24 | 1.00 (2.00) | -2.92 to 4.92 | 1.00 (-0.15 to 2.15) |
| Thrush 1994‡ | Critikon Dinamap Plus 8700 | 22 (88) | 1.41 | 1.00 (1.00) | -0.96 to 2.96 | 1.00 (-0.15 to 2.15) |
| Thrush 1994‡ | Ohmeda Oxyshuttle | 22 (88) | 3.61 | 2.00 (3.00) | -3.88 to 7.88 | 2.00 (0.85 to 3.15) |
| Thrush 1994‡ | Ohmeda Oxyshuttle | 22 (88) | 3.16 | 1.00 (3.00) | -4.88 to 6.88 | 1.00 (-0.15 to 2.15) |
| Thrush 1994‡ | Ohmeda Oxyshuttle | 22 (88) | 2.00 | 0.00 (2.00) | -3.92 to 3.92 | 0.00 (-1.15 to 1.15) |
| Thrush 1994‡ | Ohmeda Oxyshuttle | 22 (88) | 1.41 | 1.00 (1.00) | -0.96 to 2.96 | 1.00 (-0.15 to 2.15) |
| Thrush 1994‡ | MiniOx IV | 22 (88) | 3.16 | 1.00 (3.00) | -4.88 to 6.88 | 1.00 (-0.15 to 2.15) |
| Thrush 1994‡ | MiniOx IV | 22 (88) | 4.00 | 0.00 (4.00) | -7.84 to 7.84 | 0.00 (-1.15 to 1.15) |
| Thrush 1994‡ | MiniOx IV | 22 (88) | 2.24 | 1.00 (2.00) | -2.92 to 4.92 | 1.00 (-0.15 to 2.15) |
| Thrush 1994‡ | MiniOx IV | 22 (88) | 2.00 | 0.00 (2.00) | -3.92 to 3.92 | 0.00 (-1.15 to 1.15) |
| Thrush 1994‡ | Ohmeda 3700 | 22 (88) | 5.10 | 1.00 (5.00) | -8.80 to 10.80 | 1.00 (-0.15 to 2.15) |
| Thrush 1994‡ | Ohmeda 3700 | 22 (88) | 3.00 | 0.00 (3.00) | -5.88 to 5.88 | 0.00 (-1.15 to 1.15) |
| Thrush 1994‡ | Ohmeda 3700 | 22 (88) | 2.83 | 2.00 (2.00) | -1.92 to 5.92 | 2.00 (0.85 to 3.15) |
| Thrush 1994‡ | Ohmeda 3700 | 22 (88) | 2.24 | 1.00 (2.00) | -2.92 to 4.92 | 1.00 (-0.15 to 2.15) |
| Jubran 1990 | Nellcor/Ohmeda Biox 3700 | 25 (55) | 2.84 | 2.20 (1.80) | -1.30 to 5.70 | 2.20 (1.49 to 2.91) |
| Primary analysis | 13 studies (48 comparisons) | 2195 (12870) | 1.64 | 0.55 (1.55) | -2.48 to 3.58 | 0.55 (-0.21 to 1.31) |
| Sensitivity (RoB) | 8 studies (26 comparisons) | 1293 (7770) | 1.42 | 0.36 (1.38) | -2.33 to 3.06 | 0.36 (-0.88 to 1.61) |
| Sensitivity (Trans) | 10 studies (45 comparisons) | 1044 (5485) | 1.67 | 0.63 (1.54) | -2.40 to 3.65 | 0.63 (-0.38 to 1.63) |
| Sensitivity (Homo) | 8 studies (16 comparisons) | 1246 (9791) | 1.84 | 0.98 (1.56) | -2.07 to 4.04 | 0.98 (-0.08 to 2.05) |

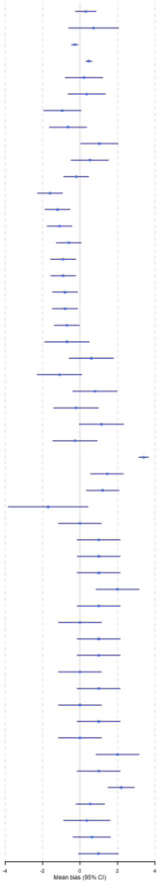

Note:

- The Chi² test for heterogeneity in the primary analysis suggested a Q(df = 47) = 1100.39, with P value < 0.0001.
- $Tau^2$ between the 13 studies = 1.10 (95% CI 0.31 to 3.48); $Tau^2$ between the 9 comparisons = 0.38 (0.24 to 0.67).
- The estimated overall $I^2$ for the primary analysis = 94.39%, of which about 69.92% is due to between-studies heterogeneity, and 24.47% due to within-study heterogeneity.

**Figure S8. Summary presentations of study sample sizes (n) and numbers of data pairs compared (N), accuracy root mean square ($A_{rms}$), mean bias (SD) and limits of agreement (LoA) of pulse oximeters for ethnic groups by the different types of pulse oximeters**

| | Studies (comparisons) | Sample (data pairs) | Arms, % | Mean bias (SD), % | LoA, % | Mean bias (95% CI), % |
|---|---|---|---|---|---|---|
| * Masimo | | | | | | |
| Black/African American | 1 study (2 comparisons) | 17 (744) | 1.90 | 0.95 (1.65) | -2.28 to 4.18 | 0.95 (-1.74 to 3.64) |
| Non-Black, non-White ethnic groups | 1 study (2 comparisons) | 7 (308) | 1.56 | 0.33 (1.53) | -2.67 to 3.32 | 0.33 (-1.69 to 2.34) |
| White/Caucasian | 1 study (2 comparisons) | 12 (526) | 1.45 | -0.22 (1.43) | -3.02 to 2.59 | -0.22 (-1.61 to 1.18) |
| * Nellcor | | | | | | |
| Black/African American | 6 studies (8 comparisons) | 202 (3658) | 2.39 | 1.81 (1.56) | -1.84 to 5.46 | 1.81 (0.57 to 3.06) |
| Non-Black, non-White ethnic groups | 2 studies (3 comparisons) | 41 (1549) | 1.20 | 0.35 (1.15) | -1.91 to 2.61 | 0.35 (-0.81 to 1.51) |
| White/Caucasian | 8 studies (20 comparisons) | 619 (8247) | 1.58 | 1.37 (1.50) | -3.52 to 4.52 | 0.50 (-0.76 to 1.76) |
| * Nonin | | | | | | |
| Black/African American | 2 studies (3 comparisons) | 24 (862) | 1.31 | 0.26 (1.29) | -2.26 to 2.78 | 0.26 (-0.73 to 1.25) |
| Non-Black, non-White ethnic groups | 1 study (2 comparisons) | 7 (308) | 1.62 | 0.27 (1.60) | -2.86 to 3.40 | 0.27 (-1.54 to 2.08) |
| White/Caucasian | 2 studies (3 comparisons) | 21 (726) | 1.27 | -0.21 (1.25) | -2.67 to 2.25 | -0.21 (-0.24 to -0.18) |
| * Philips | | | | | | |
| White/Caucasian | 1 study (2 comparisons) | 23 (138) | 1.78 | 0.70 (1.64) | -2.51 to 3.91 | 0.70 (-0.48 to 1.88) |
| * Novametrix | | | | | | |
| Black/African American | 1 study (1 comparison) | 7 (119) | 2.80 | 1.83 (2.12) | -2.33 to 5.99 | 1.83 (1.45 to 2.21) |
| White/Caucasian | 1 study (1 comparison) | 9 (200) | 2.13 | 1.15 (1.79) | -2.36 to 4.66 | 1.15 (0.90 to 1.40) |
| * Ohmeda | | | | | | |
| Black/African American | 1 study (3 comparisons) | 33 (66) | 5.42 | 5.13 (1.75) | 1.71 to 8.56 | 5.13 (0.13 to 10.13) |
| White/Caucasian | 2 studies (9 comparisons) | 30 (753) | 1.56 | -0.11 (1.55) | -4.49 to 4.27 | -0.11 (-17.00 to 16.80) |
| * Hewlett-Packard | | | | | | |
| Black/African American | 1 study (3 comparisons) | 33 (66) | 2.55 | -2.00 (1.59) | -5.11 to 1.11 | -2.00 (-5.24 to 1.24) |
| * Critikon | | | | | | |
| White/Caucasian | 1 study (4 comparisons) | 22 (352) | 1.55 | 0.75 (1.35) | -1.90 to 3.40 | 0.75 (-0.24 to 1.74) |
| * MiniOx | | | | | | |
| White/Caucasian | 1 study (4 comparisons) | 22 (352) | 1.65 | 0.50 (1.57) | -2.58 to 3.58 | 0.50 (-0.79 to 1.79) |
| * Minolta | | | | | | |
| White/Caucasian | 1 study (1 comparison) | 846 (846) | 2.38 | 0.47 (2.33) | -4.10 to 5.04 | 0.47 (0.37 to 0.57) |

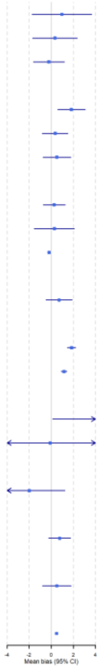

This figure presents the impact of ethnicity on pulse oximetry accuracy according to types of pulse oximeters evaluated. Results of analyses suggested that:

- Masimo, Nellcor, Novametrix, and Ohmeda appear to have higher $SpO_2$ measures than $SaO_2$ readings by on average 1% in people from Black/African American ethnic groups.
- Hewlett-Packard appears to obtain $SpO_2$ by 2% lower than $SaO_2$ readings in people with high skin pigmentation.
- Novametrix gives a $SpO_2$ measure higher than $SaO_2$ readings by on average 1% in people from White/Caucasian ethnic groups whilst others of these devices appear to produce $SpO_2$ measures with a bias no more than 1% compared with $SaO_2$ readings in White/Caucasians and those from ethnicity other than Black or White.
- Nonin does not result in over- or underestimation of oxygen saturation in people from any ethnic groups.

### Table S7. Evidence from studies that could not be included in quantitative data pooling for the ethnicity factor

| Measures of skin pigmentation | No. of participants (data pairs) and no. of studies and evaluations | Summary of reported results | Comments |
|---|---|---|---|
| ***Ethnic groups: 'moderately pigmented' or 'racially pigmented' as reported, versus unclear*** | 122 (267) in two studies with two evaluations [Escourrou 1990, Gabrielczyk 1988] | 'Racially pigmented' skin did not affect pulse oximetry accuracy:<br>• Across Ohmeda Biox 3700, Criticare CSI 501+, and Nellcor N200, the mean difference in bias between moderate pigmentation and others = 1.1% (SD 3), and skin pigmentation levels did not affect pulse oximetry accuracy.[Escourrou 1990]<br>• Nellcor N100 over-estimated oxygen saturation with a mean difference of 0.6%, and skin pigmentation levels did not affect pulse oximetry accuracy [ Gabrielczyk 1988] | **Four models evaluated*:*** Ohmeda Biox 3700, Criticare CSI 501+, Nellcor N200, Nellcor N100 |
| ***Ethnic groups: Aboriginal and/or Torres Strait Islander [ATSI] vs non-ATSI*** | 929 (18650) in one study with one evaluation [Brooks 2020] | Based on categories of mean bias > 3% vs <= 3% as the outcome measure, an univariate analysis produced OR of 0.94 (95% 0.60 to 1.48), p = 0·790, meaning ATSI was not associated with a bias higher than 3% compared with non-ATSI. Multivariate analysis produced a OR of 1.29 (95% CI 0.99 to 1.68), p = 0·055, meaning ATSI was marginally associated with a bias higher than 3% compared with non-ATSI. | **Two models evaluated together:** Masimo, Nellcor (Covidien) |
| ***Race/ethnic groups: Chinese, Malay, and Indian*** | 33 (150 readings noted, but only 98 presented) in one study with three evaluations [Lee 1993] | There was a significant difference between the groups (ANOVA, p<0.05) with the Indian group having the greatest difference between $SpO_2$ and $SaO_2$, Malay having the moderate difference, and Chinese having the least difference. | **Three models evaluated** Nellcor, Simed, Critikon |
| ***Race/ethnic groups: African American, Hispanic, White, Asian, and Other groups*** | 225 (1980) in one study with one evaluation [Ross 2014] | Based on binary categories of mean bias of < 3% and >3%, a multivariable analysis produced regression coefficients = African American -0.55 (p= .003), Hispanic -0.15 (NS), Asian -0.26 (NS), and Other -0.03 (NS), all compared with White group as the reference. A secondary analysis suggested 'a lower likelihood of bias was associated with African American race/ethnicity'. The results suggested a lower bias in African American ethnic groups than the White group. | **Three models evaluated** Masimo LCNS pulse oximeters, Nellcor oximeters (OxiMax probes), Masimo oximeters (OxiMax disposable probes) |
| ***Race/ethnic groups: Caucasian vs Black/African American*** | 43 (136) in one study with two evaluations[Schallom 2018] | • Nellcor OxiMax Forehead sensor<br>Chi-square OR (within 3% of SaO2 as the cut off): Caucasians were 1.2 times more likely to have a clinically accurate forehead measurement than African Americans. However, this association was not statistically significant (p = 0.74).<br>• Xhale Assurance nsal alar sensor<br>Chi-square OR (within 3% of SaO2 as the cut off): Caucasians were 2.65 times more likely to have a clinically accurate nasal measurement than African Americans. This association was statistically significant (p = 0.04) | **Two models evaluated** Nellcor OxiMax Forehead sensor, Xhale Assurance nsal alar sensor |
| ***Ethnic group: all Caucasian*** | 6 (NR) in one study with two evaluations [Smyth 1986] | The author fitted a regression line between $SpO_2$ and $SaO_2$. No data on mean bias was reported | **Two models evaluated** Hewlett Packard (HP) oximeter and Biox II oximeter |
| ***Ethnic group: all Chinese*** | 42 (NR) in one study with one evaluation [Stewart 1991] | The authors reported median bias of using the Ohmeda Biox 3700 model in (1) adult patients who were scheduled to undergo open heart surgery and (2) those who did not have tricuspid regurgitation by probe sites (finger and ear).<br>• Ear, patients who were scheduled to undergo open heart surgery: median (range) = 4% (0 to + 11 %),<br>• Ear, patients who did not have tricuspid regurgitation: median (range) = 1% (0 to +4%),<br>• Finger, patients who were scheduled to undergo open heart surgery: median (range) = 3% (-2 to + 10%), and<br>• Finger, patients who did not have tricuspid regurgitation: median (range) = 1% (- 1 to + 5%). | **One model evaluated** Ohmeda Biox 3700 |
